# A Systematic Literature Review of Randomized Trials Comparing In-Person and Digital Interventions for Type 2 Diabetes Prevention

**DOI:** 10.1101/2025.01.23.25321002

**Authors:** Pau Riera-Serra, David Morales-Hernández, Maria Antonia Fiol-deRoque, Rocío Zamanillo-Campos, Ignacio Ricci-Cabello

**Affiliations:** Health Research Institute of the Balearic Islands (IdISBa), 07120 Palma de Mallorca, Spain; RICAPPS- Red de Investigación Cooperativa de Atención Primaria y Promoción de la Salud - Carlos III Health Institute (ISCIII), Madrid, Spain; CIBER de Epidemiología y Salud Pública (CIBERESP), Institute of Health Carlos III, 28029 Madrid, Spain

**Author notes:** Corresponding authors: Ignacio Ricci-Cabello. Gerència d’Atenció Primària de Mallorca. Carrer de l’Escola Graduada n°3. 07003 Palma. Maria Antonia Fiol-deRoque. Gerència d’Atenció Primària de Mallorca. Carrer de l’Escola Graduada n°3. 07003 Palma.

## Abstract

**Background:** Digital and in-person lifestyle interventions to prevent type 2 diabetes (T2DM) are being increasingly implemented in some countries, particularly in the United States. However, their comparative effectiveness remains unclear, partly due to variability in intervention designs and limited robust evidence from randomized controlled trials (RCTs). Understanding their relative impacts is critical for informing evidence-based implementation in diverse healthcare settings.

**Aim:** To compare the effectiveness of digital versus in-person interventions for preventing T2DM.

**Methods:** We conducted a systematic literature review, following Cochrane methodology to identify and synthesize evidence from RCTs. Searches were conducted in EMBASE, MEDLINE, and Cochrane CENTRAL from inception to December 2024, including completed and ongoing trials published in English or Spanish. Studies comparing purely digital and in-person interventions were eligible. Meta-analyses were performed where appropriate, and narrative syntheses were provided for remaining outcomes. The GRADE approach was used to assess the certainty of evidence.

**Results:** Eight RCTs met the inclusion criteria, including six completed trials with published results and two ongoing trials. The completed trials encompassed a total of 2,450 participants across various healthcare settings. At 12 months, digital interventions were associated with significantly greater weight loss than in-person interventions (mean difference: –1.38 kg [95% CI: –2.34 to –0.43]), with moderate certainty of evidence. At shorter (3 and 6 months) and longer (>12 months) time points, no relevant differences were observed for weight, body mass index, or glycosylated haemoglobin levels between the modalities, with the certainty of evidence rated as low to very low. Evidence about cost-effectiveness was scarce. No trials evaluated key outcomes such as incidence of T2DM or health-related quality. For adverse events, no significant differences were found between modalities (RR: 1.06 [95% CI: 0.45 to 2.50]).

**Conclusions:** This systematic review highlights that while digital and in-person interventions can both be effective for T2DM prevention, their relative benefits depend on follow-up duration and contextual factors. The limited certainty of evidence and the absence of trials addressing critical outcomes, such as T2DM incidence, underscore the need for further well-designed RCTs. Future research should prioritize equivalence in intervention intensity, longer follow-up durations, and standardized reporting of outcomes to better inform public health decision-making.

## INSTRODUCTION

Type 2 diabetes mellitus (T2DM) represents one of the most significant global public health challenges of the 21st century. As of 2021, over 536 million adults worldwide were living with diabetes, with T2DM accounting for more than 90% of cases. Projections indicate that this number will rise to 783 million by 2045, driven by aging populations, urbanization, and unhealthy lifestyle behaviors[1, 2]. T2DM is a leading cause of morbidity and mortality, associated with complications such as cardiovascular disease, chronic kidney disease, and lower-limb amputations, as well as a substantial reduction in quality of life. Economically, the condition imposes a staggering burden on healthcare systems, with global health expenditures related to diabetes reaching $966 billion in 2021[1, 3].

Lifestyle interventions targeting weight loss, increased physical activity, and dietary changes have been shown to reduce the risk of developing T2DM by up to 58%, as demonstrated in landmark trials such as the U.S. Diabetes Prevention Program (DPP)[4, 5]. These programs, when effectively implemented, offer a critical strategy for alleviating the individual and societal burden of T2DM. However, scaling these interventions to diverse populations and healthcare settings remains a challenge.

In-person delivery of lifestyle interventions, typically conducted through structured group sessions led by trained professionals, has demonstrated high levels of engagement and efficacy in clinical trials[5, 6]. However, this modality can be resource-intensive, requiring significant investments in trained personnel, infrastructure, and participant time, which may limit its scalability and accessibility[7]. In contrast, digital interventions leverage technologies such as mobile applications, wearable devices, and online platforms to deliver content and support remotely. These approaches are cost-effective, scalable, and more flexible for participants, but they often face challenges such as lower adherence rates and variability in digital literacy[8, 9]. Both modalities have shown promise in reducing diabetes risk, but their comparative effectiveness in real-world settings remain underexplored.

Previous studies addressing this question have often relied on observational designs or indirect comparisons. For instance, several cohort studies have examined the outcomes of DPPs implemented at scale, comparing digital and in-person interventions within separate cohorts[10]. Although these studies provide valuable insights, they are prone to confounding and selection bias, which limit their ability to establish causality. Similarly, meta-analyses that aggregate data from heterogeneous studies often lack the granularity needed to isolate differences between delivery modalities[11, 12]. Randomized controlled trials (RCTs) offer the highest level of evidence, minimizing biases through randomization and enabling direct comparisons.

The objective of this review is to synthesize evidence on the comparative effectiveness of in-person versus digital interventions for preventing T2DM, thereby providing insights to guide the design and implementation of effective and equitable diabetes prevention strategies.

## METHODS

This systematic review has been conducted following the methods from Cochrane Collaboration for systematic reviews of interventions[13]. The methods and findings of this manuscript are reported following the Preferred Reporting Items for Systematic Reviews and Meta-Analysis (PRISMA) statement[14].

### Data sources and search methodology

We searched for randomized trials in MEDLINE (Ovid), EMBASE (EMBASE), and Cochrane Central Register of Controlled Trials (CENTRAL, Cochrane Library) using predefined search strategies from inception to December 2024. The search strategies (available in Appendix 1) combined MeSH terms and keywords.

### Study selection

We applied the following inclusion criteria:

#### Population

Prediabetic adult population (i.e., people with high risk of developing T2DM). Any definition of prediabetes was accepted.

#### Intervention

T2DM prevention intervention fully delivered remotely (digitally delivered).

#### Comparator

T2DM prevention intervention fully delivered in-person.

#### Outcomes

This systematic review included studies that evaluated the impact of interventions on several outcomes identified and prioritized by an international expert panel comprising clinicians, academics, and patient representatives (see Acknowledgements). The panel evaluated a comprehensive list of 28 potential outcomes, rating each as critical (2 points), important (1 point), or not important (0 points) based on clinical relevance and patient-centered considerations. Mean scores were calculated and rescaled to a 0–10 scale (refer to Online Appendix 2). We included studies that evaluated at least one of the following outcomes: three outcomes classified as critical (mean score: 9 –10): body mass index (BMI), glycosylated hemoglobin (HbA1c), and quality of life; and nine outcomes classified as important (mean score: 7–8): waist circumference, physical activity level, body weight, diet, incidence of T2DM, plasma glucose levels (2-hour oral glucose tolerance test), sedentary behavior, self-reported smoking behavior, and cost-effectiveness.

We included randomized trials, (including cluster randomized trials). Trial protocols were also included. Due to constrained resources, we only included studies published in English or Spanish. Interventions combining in-person and remote delivery were excluded. We also excluded conference abstracts, letters to the editor, and editorials.

One author (IRC) screened the search results based on title and abstract. Two authors (IRC and DMH) independently assessed eligibility based on the full text of the relevant articles. Disagreements were discussed (involving a third author when needed) until consensus was reached.

### Data extraction and risk of bias assessment

From each primary study, data on its study design, population, setting, follow-up, results (including adverse events) were independently extracted by one reviewer (DMH) using an *ad hoc* data extraction form which had been piloted in advance. A second author (IRC, MFdR, PRS) cross-checked the extracted data for accuracy.

We applied the Cochrane RoB tool2.0 [15] to determine the risk of bias of the included trials. The ROB was judged as “low”, “some concerns”, or “high risk” for each of the following domains: randomization sequence; deviation from intended interventions; missing outcome data; measurement of the outcome, and; selection of the reported results. Risk of bias was measured at the outcome (rather than at the trial) level. One author determined the risk of bias of the included studies (PRS), and a second author cross-checked the results for accuracy (DMH). Disagreements were solved with support from a senior systematic reviewer (IRC).

### Data synthesis

We described the interventions’ characteristics and effects narratively and as tabulated summaries. Findings are summarized by follow-up period (3 months, 6 months, 12 months, and beyond 12 months).

When appropriate data was available, we performed a random-effects meta-analysis using the DerSimonian & Laird method. For continuous outcomes, we performed a meta-analysis on the change from baseline values. Statistical heterogeneity was assessed with the Q-Cochran test and with the *I*^2^ statistic[16]. A *p*-value of the Q-Cochran test <0.10 and an *I*^2^>50% were considered to indicate substantial heterogeneity. All statistical analyses were performed using STATA software, v.15.

### Certainty of the evidence

We rated the certainty (quality) of evidence for each outcome using the GRADE approach [17]. Certainty of evidence was rated as “high”, “moderate”, “low” or “very low”, taking into consideration the risk of bias, imprecision, inconsistency, indirectness, and publication bias domains. Results were summarised in summary of findings tables [18]. One author graded the evidence of included studies (IRC), and a second author cross-checked the results for accuracy (PRS). Disagreements were solved with support from a third reviewer (MFdR).

## RESULTS

### Selection process

The eligibility process is summarized in a PRISMA flowchart (Fig. 1). We retrieved a total of 1,304 unique citations from database searches. We selected 50 references for full text revision, from which 16 publications [19–34] (reporting on 8 different trials) were finally included in our systematic review. The reason for exclusion at full-text level are detailed in Online Appendix 3. Two [19, 23, 31]of the eight trials had not been completed, and their results were unavailable at the time the searches were conducted (Online Appendix 4).

**Figure 1.**
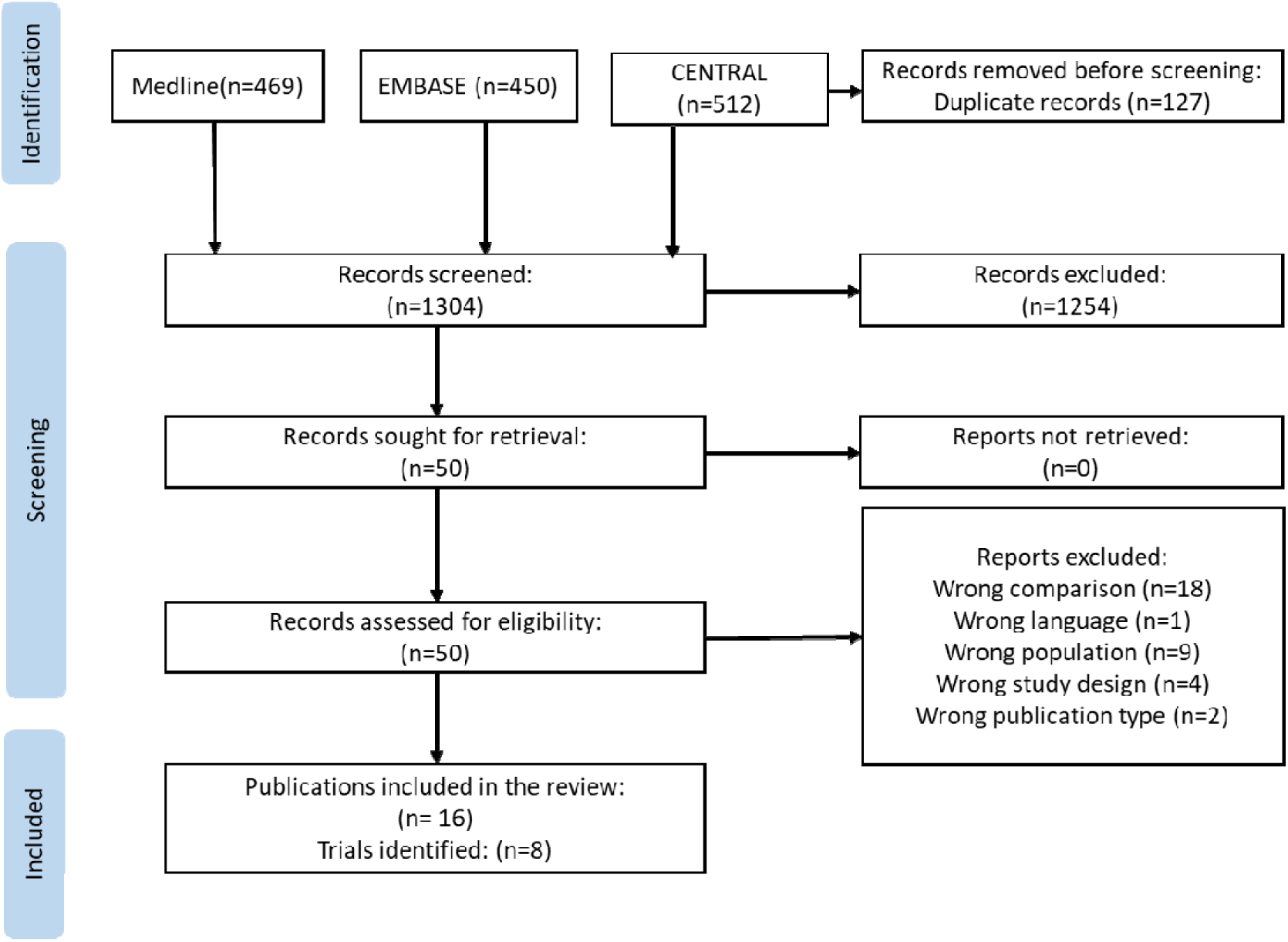
PRISMA flowchart

### Characteristics of the included trials

The characteristics of the six completed trials are described in Table 1. The trials were conducted in diverse settings, predominantly in the United States (five trials) and one in the Czech Republic[29]. The trial designs varied, including parallel-group randomized controlled trials [19, 20, 23–32, 34–36], and one hybrid preference-randomized design [21, 22]. Follow-up durations ranged from 6 months to 18 months. The sample sizes across trials varied significantly, with the smallest trial enrolling 100 participants [21, 22] and the largest involving 482 participants [25]. Most studies achieved a balanced or moderately higher proportion of female participants, with gender representation ranging from 47% to 71%. Participant age also varied, with mean ages ranging from 43.3 to 52.9 years.

**Table 1.** Characteristics of the completed trials identified.

| Trial name/<br>Trial<br>registration<br>code | Country/<br>number of<br>participants | Trial design | Inclusion criteria | Trial<br>follow-up | Proportion<br>female | Mean (SD)<br>age | Description of in-person<br>intervention | Description of digital intervention |
| --- | --- | --- | --- | --- | --- | --- | --- | --- |
| CCSF<br>Diabetes<br>Prevention<br>Trial<br>(Ferrara<br>2020)[24]/<br>not<br>reported | US/ 158 | 2-arm, parallel,<br>randomized<br>controlled trial | CCSF employees, CDC<br>prediabetes screener score $\geq 9$,<br>BMI $\geq 25$ kg/m<sup>2</sup> ($\geq 23$ kg/m<sup>2</sup> if<br>Asian); prediabetes defined by<br>high-risk screener score; no<br>major health conditions;<br>regular computer/internet<br>access; not in weight loss<br>programs. | 6-month<br>and 12-month<br>follow up | 56% | 52.7 (8.7)<br>in-person,<br>51.8 (7.8)<br>digital | YMCA-DPP: in-person<br>intervention with 16 weekly 1-hour sessions, 3 biweekly, and 5<br>monthly sessions, conducted for<br>groups of 8-15 employees at<br>convenient worksite times. | VLM-DPP: online intervention via<br>Canary Health with 16 weekly<br>educational sessions and 8 monthly<br>maintenance sessions,<br>supplemented by a single in-person<br>orientation. Sessions (25-45 min)<br>included audio and interactive<br>visuals. |
| DiaBEAT-it[21, 22]/<br>NCT021629<br>01 | US/ 334 | Hybrid 2-group<br>preference<br>(Choice) and 3-group randomized<br>controlled | Age $\geq 18$, BMI $\geq 25$ kg/m<sup>2</sup> ($\geq 22$ if<br>Asian), prediabetes DRC test<br>score $\geq 5$, English-speaking, not<br>pregnant, no T2D or major<br>heart conditions, no physical<br>activity/weight loss<br>contraindications, phone<br>access. | 6, 12, and<br>18<br>months | 68% | 52,3 (12,1) | 2-hour small group class focused<br>on developing a personal action<br>plan for T2D prevention,<br>targeting 10% weight loss and 60<br>minutes of physical activity, 5<br>days/week, with guidance on diet<br>and exercise. | DVD-based intervention (60 min)<br>with 22 tailored IVR calls over 12<br>months, covering pre-diabetes, risk<br>factors, action plans, goal setting,<br>and behaviour change, targeting<br>10% weight loss and 60 min of<br>activity, 5 days/week. |
| E-LITE[26]/<br>NCT008424<br>26 | US/241 | 3-arm, parallel,<br>randomized<br>controlled trial (digital<br>intervention, in-person<br>intervention, and<br>usual care) | Age $\geq 18$, BMI $\geq 25$, prediabetes<br>(fasting glucose 100–125<br>mg/dL) or metabolic syndrome<br>(AHA/NHLBI criteria). | 3, 6, and<br>15<br>months | 47% | 52.9 (10.6) | 3 months of intensive<br>intervention (12-session GLB<br>curriculum) followed by 12<br>months of maintenance. Sessions<br>included food tastings, physical<br>activity, and were led by a<br>dietitian and fitness instructor. | 3 months of intensive intervention<br>(12-session GLB curriculum) and 12<br>months of maintenance.<br>Participants used a home-based<br>DVD, attended orientation, and<br>used Heart360 for goal setting and<br>monitoring |
| Fuel Your<br>Life[32, 34]/<br>not<br>reported | US/418 | 3-arm, parallel,<br>randomized<br>controlled trial (1)<br>self-study (no<br>coaching), (2)<br>small-group<br>facilitated by a | City-county government<br>employees; all eligible,<br>recruitment targeted to high-risk diabetes (Diabetes Risk<br>Screener high-risk score or BMI<br>$\geq 30$). | 6 months<br>// 3<br>months<br>(midpoint)<br>, 6<br>months,<br>and 12 | 59% in-person,<br>64% digital<br>// 58,2%<br>in-person,<br>67,9%<br>digital | 47 // 45.91<br>in-person,<br>47.75<br>digital | Group coaching: 50-minute<br>sessions at worksites with 5-10<br>participants. Small groups of 8-10<br>met biweekly for 4 sessions,<br>monthly for 4 sessions, and bi-monthly during a 6-month<br>maintenance phase. | Phone sessions: 20-minute one-on-one telephone sessions with a<br>health coach, offering tailored<br>support and accountability, but<br>without group reinforcement or<br>support, following the same<br>schedule of in-person (group |
|  |  | health coach (group coaching), and (3) individual health coaching by phone (phone coaching). |  | months (follow-up) |  |  |  | coaching) sessions |
| Moravcova 2024[29]/ NCT04573296 | Czech Republic/100 | prospective, double-armed, randomized non-inferiority trial | Age ≥18, BMI >30, diagnosed with T2D, prediabetes (fasting glucose 5.6–6.9 mmol/L, OGTT 7.8–11.0 mmol/L), or insulin resistance (HOMA-IR >2.7), smartphone for Vitadio app, able to comply with study procedures | 6 months | 71% | 43.3 ± 9.5 | Five in-person lifestyle consultations with a physician, dietitian, or nurse, focusing on healthy eating, balanced diets (fruits, vegetables, healthy fats, whole grains, lean proteins), and an online food diary for tracking. | Vitadio: digital care program with a 3-month intensive phase followed by a sustaining phase, utilizing personalized daily tasks and automated messages to help patients establish a healthy routine. |
| PREDICTS[20, 25, 27, 28, 30, 33]/ NCT03312764 | US/482 | single-blind randomized controlled trial (RCT) design | Age ≥19, BMI ≥25 kg/m<sup>2</sup> (≥22 if Asian), HbA1c 5.7%–6.4%, medically stable (no uncontrolled hypertension, hyperlipidemia, cancer treatment, untreated metabolic condition), not a bariatric surgery candidate, capable of consent, email/internet access | 4 and 12 months | not reported | ≥ 19 | 2-hour small group session focusing on action planning for increased physical activity, improved nutrition, weight loss, goal setting, and barrier resolution, with a resource booklet on local lifestyle support. | Internet-based intervention with small group support, personalized health coaching, digital tracking tools, and weekly behaviour change curriculum, approved by the Diabetes Prevention Recognition Program. |

The trials used various methods to define and measure prediabetes. The CCSF Diabetes Prevention Trial[24] relied on a CDC prediabetes screener with a high-risk score (≥9). The DiaBEAT-it trial[21, 22] used a Diabetes Risk Calculator (DRC) test score of ≥5. The E-LITE trial[26] defined prediabetes by fasting plasma glucose levels of 100–125 mg/dL or the presence of metabolic syndrome based on AHA/NHLBI criteria. The Fuel Your Life trial[32, 34] targeted participants with a high-risk Diabetes Risk Screener score or a BMI ≥30. The trial by Moravcová et al[29] defined prediabetes as fasting glucose between 5.6–6.9 mmol/L or OGTT results of 7.8–11.0 mmol/L. Lastly, the PREDICTS trial[27] identified prediabetes by baseline HbA1c levels between 5.7% and 6.4%.

### Characteristics of the digital and in-person interventions compared by the trials

In the CCSF Diabetes Prevention Trial[24], the in-person intervention consisted of 16 weekly group sessions lasting one hour each (core curriculum) followed by 3 biweekly and 5 monthly sessions (maintenance curriculum). These sessions were conducted at employees’ worksites, accommodating their schedules during breaks or lunch hours, with groups of 8 to 15 participants. The digital intervention consisted in 16 weekly educational sessions and 8 monthly maintenance sessions, supplemented by a single in-person orientation. Sessions (25- 45 min) included audio and interactive visuals.

In the DiaBEAT-it trial[21, 22], the in-person intervention consisted in a 2-hour in-person small group class focused on helping participants create a personalized action plan to prevent diabetes, including setting a goal to lose 10% of their body weight within 12 months and engaging in 60 minutes of physical activity five days per week. Instructors provided detailed information on healthy eating and physical activity based on MyPlate guidelines and distributed workbooks covering 22 educational topics adapted from the original DPP. Thecomprised a 60-minute educational DVD with segments addressing prediabetes, diabetes risk factors, action plan development, goal setting for physical activity and nutrition, resource building, and commitment to change. Additionally, participants in the digital intervention received 22 personalized IVR calls over 12 months, designed to reinforce weight loss and healthy behavior maintenance.

In the E-LITE trial [26], the in-person intervention consisted in 3 months of intensive intervention (12-session GLB curriculum) followed by 12 months of maintenance. Sessions included food tastings, physical activity, and were led by a dietitian and fitness instructor. The digital intervention was a self-directed approach utilizing DVDs that delivered the same DPP-based content. Participants engaged with the material independently, receiving guidance on implementing lifestyle changes to facilitate weight loss and improve health outcomes.

In the “Fuel Your Life” trial, the in-person intervention small groups of 8-10 met biweekly for 4 sessions, monthly for 4 sessions, and bi-monthly during a 6-month maintenance phase where participants engaged in discussions and activities led by a facilitator, focusing on diet, physical activity, and behavior change strategies. The digital intervention consisted of a telephone intervention. The telephone sessions were held one-on-one between the health coach and the participant, following the same schedule as the in-person intervention. Each session lasted around 20 minutes, with the coach calling the participant at a prearranged time using a contact number provided by the participant. While the coach could better customize the discussion and actions to the individual, they were solely responsible for providing reinforcement and accountability, as there was no group support.

In the trial conducted Moravcová et al. (2024)[29], the in-person intervention comprised five regular sessions with a physician, dietitian, or nurse focusing on lifestyle modifications, including dietary changes and physical activity. The digital intervention utilized the Vitadio application, a digital care program with a 3-month intensive phase followed by a sustaining phase, utilizing personalized daily tasks and automated messages to help patients establish a healthy routine

In the PREDICTS trial[20, 25, 27, 28, 30, 33], the in-person consisted of a 2-hour small group session focusing on action planning for increased physical activity, improved nutrition, weight loss, goal setting, and barrier resolution, with a resource booklet on local lifestyle support. The digital intervention consisted of an Internet-based intervention with small group support, personalized health coaching, digital tracking tools, and weekly behaviour change curriculum, approved by the Diabetes Prevention Recognition Program.

### Risk of bias

A description of the risk of bias of the included trials is depicted in Figure 2. Overall, one trial (PREDICTS)[20, 25, 27, 28, 30, 33] was rated as “low risk”. Four trials were classified as having “some concerns,” primarily due to issues with randomization, protocol transparency, and handling of missing data. One trial was judged as having “high risk” due to a general lack of specific details regarding randomization, blinding, allocation process and protocol availability

**Figure 2.**
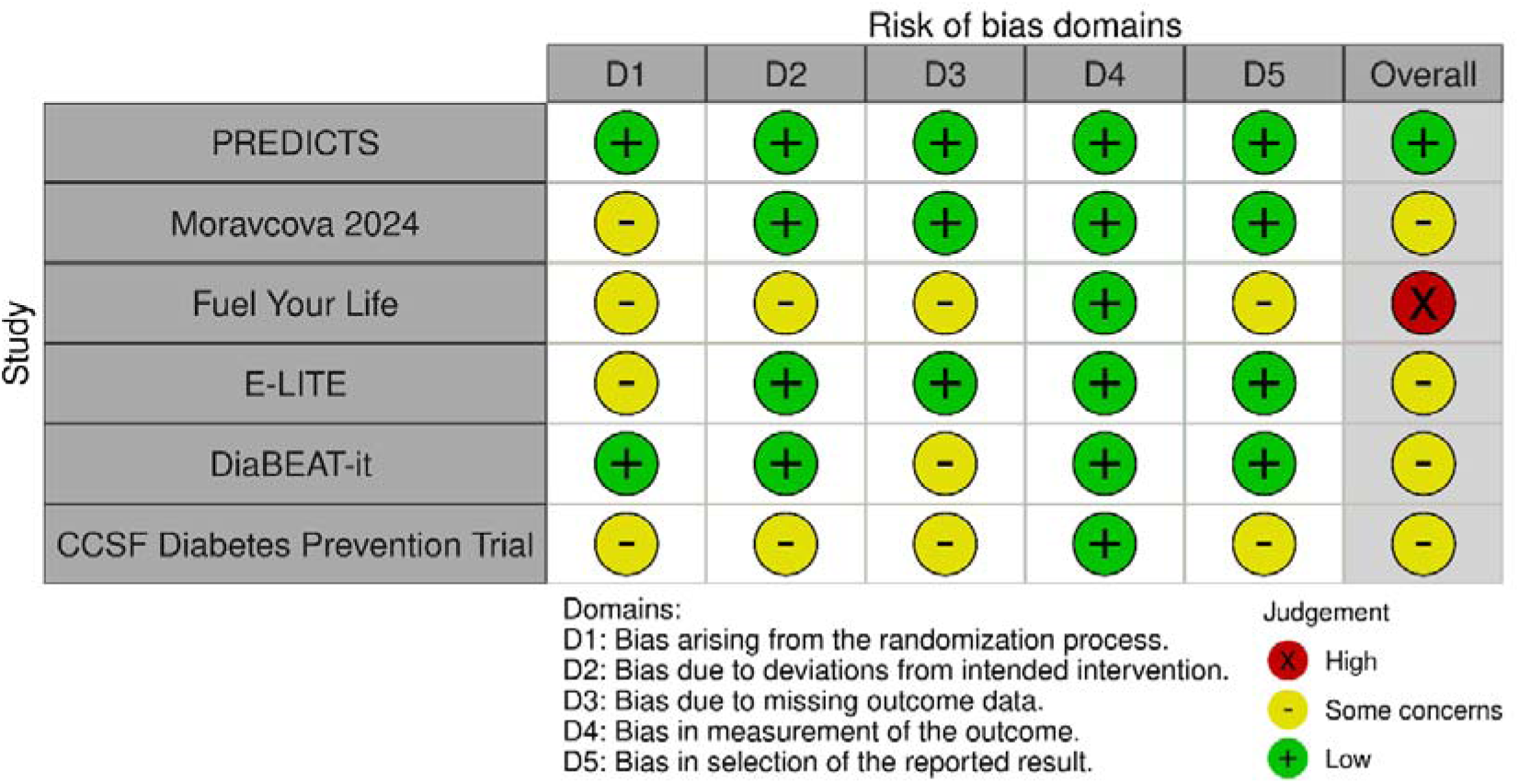
Risk of bias of the included trials

For the randomization process, two trials (PREDICTS and DiaBEAT-it) were assessed as “low risk,” while the remaining four trials were rated as having “some concerns” due to incomplete or unclear information regarding allocation sequence and concealment methods. Regarding deviations from intended interventions, four trials (PREDICTS, Moravcová 2024, E-LITE, and DiaBEAT-it) were rated as “low risk,” while the others (CCSF Diabetes Prevention Trial and Fuel Your Life) were assigned “some concerns,” primarily due to lack of information about blinding or adherence to the intervention protocol. For missing outcome data, three trials (PREDICTS, Moravcová 2024, E-LITE) were rated as “low risk,” and three (Fuel Your Life, DiaBEAT-it and and CCSF Diabetes Prevention Trial) as having “some concerns,” mostly due to high dropout rates or insufficient justification for handling missing data. Measurement of the outcome was rated as “low risk” in all six trials, indicating robust and unbiased outcome assessment methods. For selection of the reported result, four trials (PREDICTS, E-LITE, DiaBEAT-it, and Moravcová 2024) were rated as “low risk,” while the remaining two (Fuel Your Life and CCSF Diabetes Prevention Trial) were rated as having “some concerns,” reflecting a lack of protocol availability or inconsistencies in reporting.

### Comparison of the effects of digital vs in person interventions at three months

Table 2 presents a comparison of the effects of digital versus in-person interventions on the outcomes prioritized by the expert panel. The findings are based on the meta-analyses conducted at three months of follow-up and are accompanied by an assessment of the certainty of evidence using the GRADE approach.

**Table 2.**
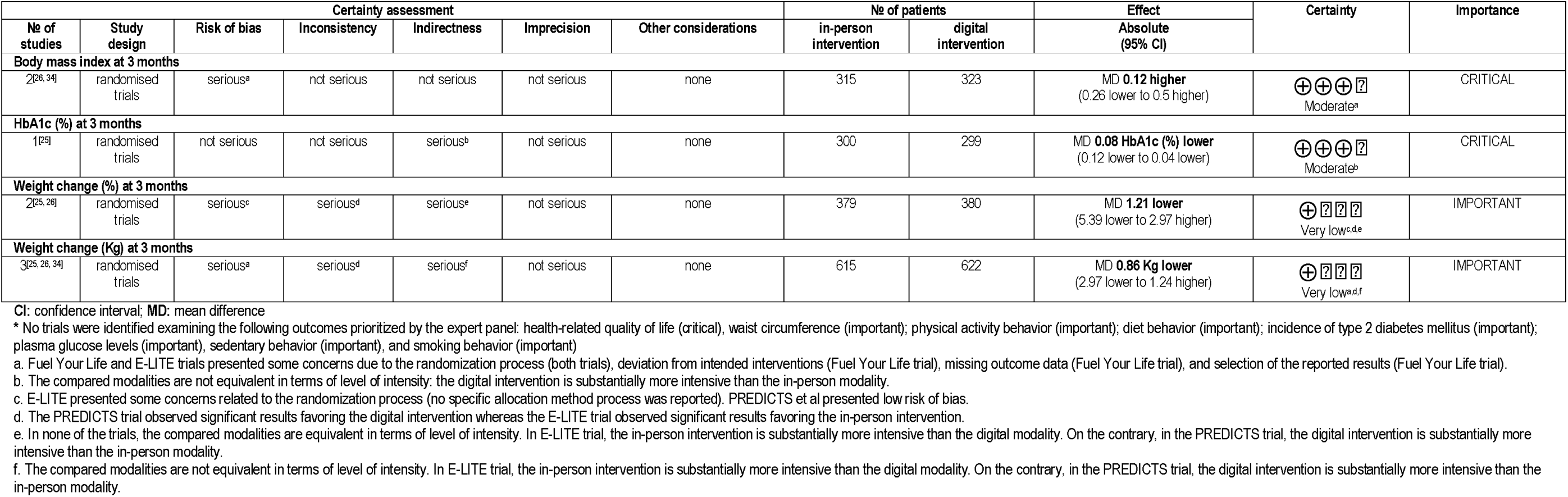
GRADE evidence and summary of findings of the comparison of the effects at three months follow-up of digital vs in person interventions to prevent type 2 diabetes mellitus*

Body mass index: Two trials [26, 34] assessed the impact of digital versus in-person interventions on BMI at three months. The meta-analysis (Online Appendix 6) showed no significant differences between the two modalities (mean difference: 0.12 higher [–0.26 to 0.50]). The certainty of evidence was rated as moderate due to serious risk of bias.

HbA1c: One trial [25] assessed changes in HbA1c at three months. The digital intervention showed a small reduction in HbA1c compared to the in-person modality (mean difference: – 0.08% [–0.12 to –0.04]). The certainty of the evidence was rated as moderate due to serious indirectness.

Body weight: Two trials [25, 26] evaluated percentage weight change at three months. The pooled analysis indicated no significant differences between digital and in-person interventions (mean difference: –1.21% [–5.39 to 2.97]). The certainty of the evidence was rated as very low due to serious risk of bias, inconsistency across trials, and indirectness. Three trials [25, 26, 34] analysed weight change in kilograms at three months. The meta-analysis found no significant differences between the modalities (mean difference: –0.86 kg [–2.97 to 1.24]). The certainty of the evidence was rated as very low due to serious risk of bias, inconsistency, and indirectness.

### Comparison of the effects of digital vs in person interventions at six months

Body mass index: Three trials [21, 22, 26, 34] assessed the impact of digital versus in-person interventions on BMI at six months. The pooled analysis indicated no significant differences between the two modalities (mean difference: –0.02 [–1.13 to 1.1]). The certainty of evidence was rated as low due to serious risk of bias and inconsistency across the trials (Table 3).

**Table 3.**
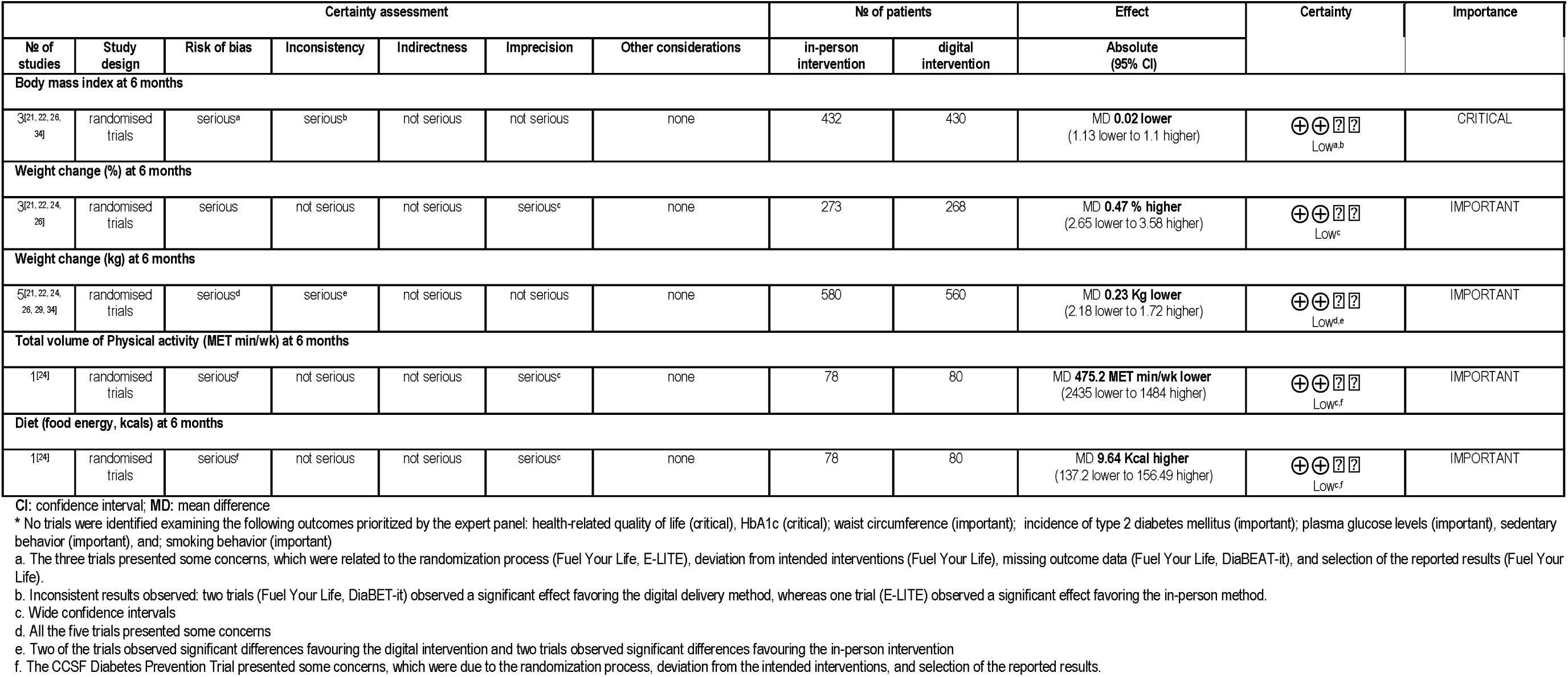
GRADE evidence and summary of findings of the comparison of the effects at six months follow-up of digital vs in person interventions to prevent type 2 diabetes mellitus*

Body weight: Three trials [21, 22, 24, 26] evaluated percentage weight change at six months. The meta-analysis showed no significant difference between the modalities (mean difference: 0.47% higher [–2.65 to 3.58]). The certainty of the evidence was rated as low, due to serious concerns related to risk of bias and imprecision. Five trials [21, 22, 24, 26, 29, 34] examined weight change in kilograms at six months. The pooled results showed no significant differences between digital and in-person interventions (mean difference: –0.23 kg [–2.18 to 1.72]). The certainty of the evidence was rated as low due to serious risk of bias and inconsistency.

Physical activity: One trial [24] evaluated the total volume of physical activity at six months, observing no significant differences between the in-person and digital interventions (mean difference: 475 MET min/week [-2435 to 1484]). The certainty of the evidence was rated as low due to serious risk of bias and imprecision.

Diet: One trial [24] assessed the impact of digital versus in-person interventions on food energy at six months, observing no significant differences (mean difference: 9.64 kcal [-137.2 to 156.5]). The certainty of the evidence was rated as low due to serious risk of bias and imprecision.

### Comparison of the effects of digital vs in person interventions at 12 months

Body mass index: Two trials [21, 22, 34] evaluated the impact of digital versus in-person interventions on BMI at 12 months. The pooled analysis showed that the digital intervention resulted in a slightly lower BMI compared to the in-person modality (mean difference: –0.5 [– 0.6 to –0.39]). The certainty of evidence was rated as moderate due to serious risk of bias (Table 4).

**Table 4.**
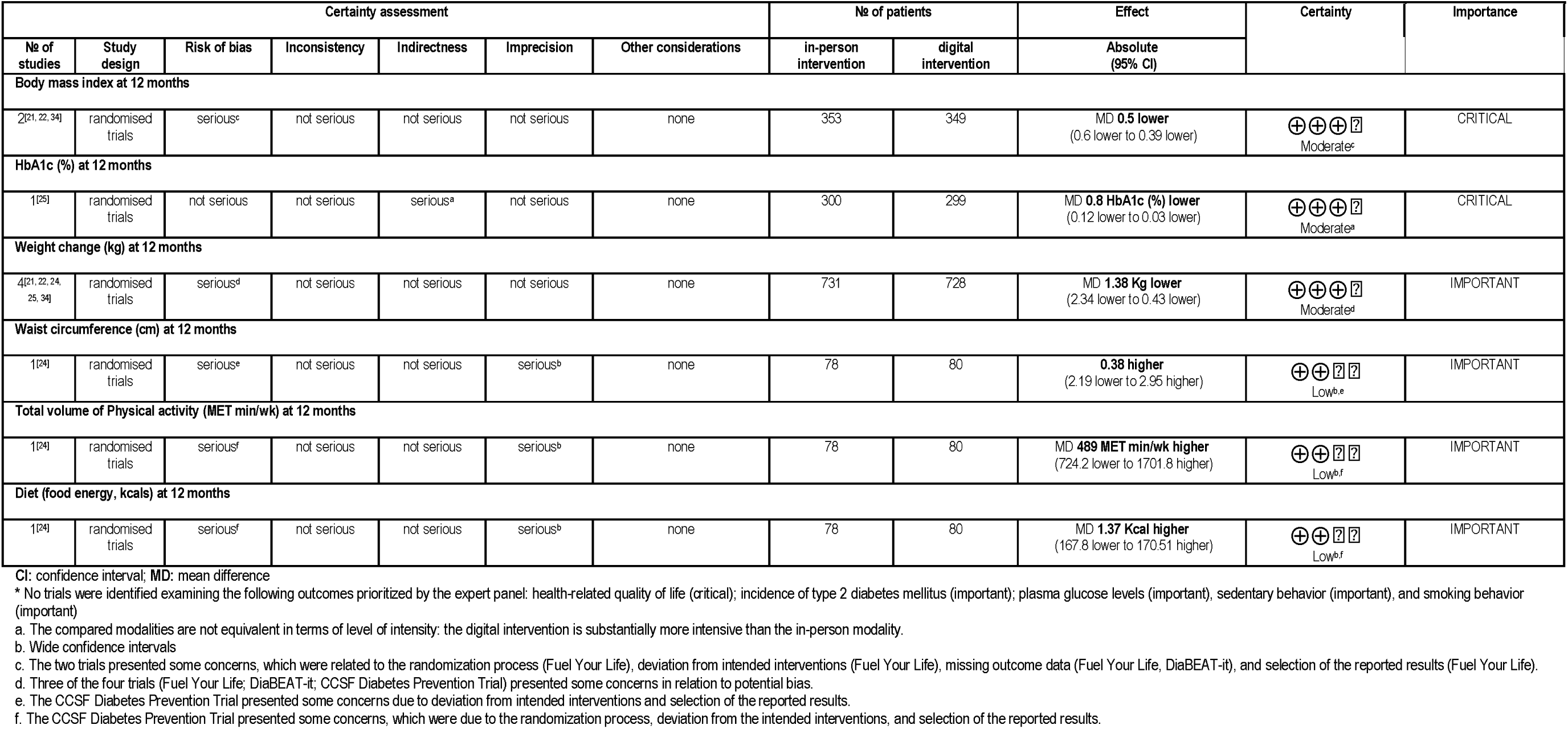
GRADE evidence and summary of findings of the comparison of the effects at 12 months follow-up of digital vs in person interventions to prevent type 2 diabetes mellitus*

HbA1c: One trial [25] assessed changes in HbA1c at 12 months. The digital intervention was associated with a small reduction in HbA1c compared to the in-person modality (mean difference: –0.08% [–0.12 to –0.03]). The certainty of evidence was rated as moderate due to serious indirectness, as the compared modalities differed significantly in intensity.

Body weight: Four trials [21, 22, 24, 25, 34] analysed weight change in kilograms at 12 months. The meta-analysis indicated that the digital intervention was associated with slightly greater weight loss compared to the in-person intervention (mean difference: –1.38 kg [–2.34 to – 0.43]). The certainty of the evidence was rated as moderate, primarily due to concerns about risk of bias.

Waist circumference: One trial [24] evaluated waist circumference at 12 months. The analysis showed no significant difference between the modalities (mean difference: 0.38 cm higher [–2.19 to 2.95]). The certainty of evidence was rated as low, due to wide confidence intervals and risk of bias.

Physical activity: One trial [24] evaluated the total volume of physical activity at 12 months, observing no significant differences between the in-person and digital interventions (mean difference: 489 MET min/week [-724 to 1701]). The certainty of the evidence was rated as low due to serious risk of bias and imprecision.

Diet: One trial [24] assessed the impact of digital versus in-person interventions on food energy at 12 months, observing no significant differences (mean difference: 1.37 kcal [-167.8 to 170.1]). The certainty of the evidence was rated as low due to serious risk of bias and imprecision.

### Comparison of the effects of digital vs in person interventions at more than 12 months

Body mass index: Two trials [21, 22, 26] assessed the effects of digital versus in-person interventions on BMI at more than 12 months. The pooled analysis showed no significant differences between the modalities (mean difference: 0.00 [–1.18 to 1.18]). The certainty of evidence was rated as very low due to serious risk of bias and inconsistency across trials (Table 5).

**Table 5.**
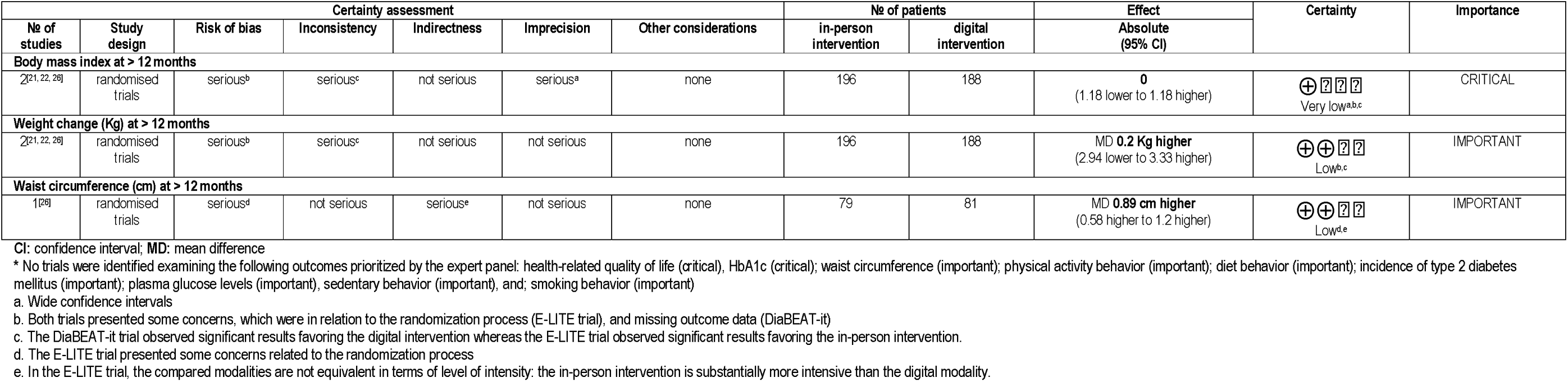
GRADE evidence and summary of findings of the comparison of the effects at >12 months follow-up of digital vs in person interventions to prevent type 2 diabetes mellitus*

Body weight: Two trials [21, 22, 26] evaluated weight change in kilograms at more than 12 months. The meta-analysis indicated no significant difference between digital and in-person interventions (mean difference: 0.2 kg [–2.94 to 3.33]). The certainty of the evidence was rated as low, primarily due to concerns about risk of bias and inconsistency.

Waist circumference: One trial [24] assessed waist circumference at more than 12 months. The analysis showed no significant differences between digital and in-person interventions (mean difference: –0.42 cm [–2.20 to 1.36]). The certainty of evidence was rated as low due to serious concerns about imprecision and bias.

### Comparison of the adverse events of digital vs in person interventions

Three trials [22, 25, 26] compared adverse events associated with digital and in-person interventions for T2DM prevention, with pooled results showing no significant differences between the modalities (iRR 1.06 [95% CI: 0.45 to 2.50]). Most reported adverse events were minor, such as musculoskeletal discomfort related to increased physical activity, and no serious adverse events attributable to either intervention were observed. However, the certainty of the evidence was rated as low due to inconsistent reporting across trials, variability in how adverse events were monitored, and imprecision stemming from the small number of studies.

### Comparison of the cost-effectiveness of digital vs in person interventions

Cost-effectiveness was evaluated only in the PREDICTS trial [28, 33], which compared digital and in-person interventions from both private payer and societal perspectives. The digital intervention was slightly more expensive than the in-person modality from a private payer perspective ($4,556 vs. $4,177) but less expensive from a societal perspective ($13,093 vs. $14,731). In terms of health benefits, the difference in quality-adjusted life years (QALYs) between the modalities was minimal (0.019 QALYs; 95% CI: −0.003 to 0.040). The digital intervention was considered more cost-effective at willingness-to-pay (WTP) thresholds of $50,000, $100,000, and $150,000 per QALY gained.

## DISCUSION

### Main findings

This systematic review included eight RCTs comparing purely digital and in-person interventions for T2DM prevention, with six completed trials involving 2,450 participants. At 12 months, digital interventions showed significantly greater weight loss (mean difference: –1.38 kg [95% CI: –2.34 to –0.43]), though evidence certainty was moderate. At 3, 6, and more than 12 months, no relevant differences were observed for BMI, HbA1c, or weight, with consistently low to very low certainty. No trials assessed outcomes rated as critical, like glycosylated hemoglobin (HbA1c) or health-related quality of life.

### Strengths and limitations

This systematic review has several strengths. Conducted using Cochrane methodology, it ensures transparency, reproducibility, and adherence to best practices. Comprehensive searches in EMBASE, MEDLINE, and Cochrane CENTRAL identified both completed and ongoing trials. By including only randomized controlled trials comparing purely digital and in-person interventions, the review minimizes bias and ensures internal validity. Outcomes were assessed at multiple follow-up points, providing insights into the sustainability of effects. Risk of bias was systematically evaluated, and outcomes prioritized by experts align with clinical and patient-centred priorities.

This systematic review has several limitations that should be acknowledged. First, while comprehensive searches were conducted across major databases, excluding grey literature and non-indexed sources might have omitted studies with valuable insights, particularly unpublished or ongoing research. Second, the inclusion of studies was restricted to those published in English or Spanish, potentially excluding relevant evidence available in other languages. Third, the limited number of randomized controlled trials identified reduced the robustness of publication bias assessments, as statistical tools for detecting such bias require a larger pool of studies to yield reliable results. Additionally, the inclusion criteria, while designed to ensure a focused comparison between purely digital and purely in-person interventions, excluded hybrid models that are increasingly relevant in real-world practice. This could limit the generalizability of the findings to mixed-delivery approaches, which are becoming more common. Finally, despite efforts to compare equivalent delivery modalities, differences in the intensity, structure, and duration of interventions introduce heterogeneity that complicates direct comparisons.

### Comparison with previous literature

Our systematic review offers novel evidence due to its focus on clinical trials directly comparing purely digital and purely in-person interventions for T2DM prevention. This approach allows for a higher level of experimental evidence compared to the observational studies that dominate much of the existing literature. However, for most outcomes examined, the certainty of evidence was rated as low or very low, and key outcomes prioritized by our expert panel, such as HbA1c or health-related quality of life, were not assessed in the available trials.

Observational studies, while limited by potential confounding, offer complementary insights into the effectiveness of digital and in-person interventions. For example, Ely et al. (2023) [37] and Villegas et al. (2022) [38] found that digital interventions can achieve weight loss outcomes comparable to in-person modalities, particularly when participants are allowed to choose their preferred delivery method. Conversely, Bian et al. (2017) [39], observed slightly lower weight loss with digital interventions, which was attributed to challenges in engagement and accountability. These mixed findings reflect the variability in intervention designs and the contextual factors influencing their effectiveness.

Our review contributes to this body of evidence by identifying a statistically significant greater weight loss with digital interventions compared to in-person modalities at 12 months. However, this finding must be interpreted considering the low certainty of evidence and the lack of standardized intervention designs across trials. At shorter follow-up periods (3 and 6 months), as well as at longer follow-up periods (>12 months) no significant differences between the two modalities were observed for most outcomes (with low or very low certainty), further emphasizing the need for additional high-quality trials with rigorous designs and long-term follow-up.

### Key evidence gaps and future research needs

Despite advances in understanding the comparative effectiveness of digital and in-person interventions for T2DM prevention, significant gaps in the evidence remain. A notable limitation is the lack of equivalence between the interventions being compared in existing trials. Digital and in-person interventions often differ significantly in intensity, structure, and participant engagement, making it challenging to isolate the true impact of the delivery modality. For example, in some trials, in-person interventions involved structured group sessions with real-time interaction, while digital interventions relied on self-paced or asynchronous formats with limited feedback. Future research should prioritize designing interventions that are equivalent in content, intensity, and support mechanisms to enable more accurate comparisons.

A notable gap is the geographic concentration of evidence, with most trials, including those currently underway (Abusamaan 2023[19, 31]; Beasley 2023[23]), conducted in the United States. Expanding research to other regions, such as Europe, where both modalities are implemented, is essential to improve generalizability. Additionally, key outcomes prioritized by expert panels, such as quality of life and HbA1c, are underreported. Only the PREDICTS trial evaluated cost-effectiveness, and few studies included long-term follow-up data, limiting insights into the sustainability of intervention effects. There is also variability in how outcomes like weight loss and physical activity are measured across trials, complicating meta-analyses. Standardized metrics and protocols are needed to enhance comparability.

Finally, the evidence lacks diversity in studied populations, with limited inclusion of rural, low-income, or older adult populations. Few studies address scalability factors, such as implementation fidelity, acceptability, or barriers in low-resource settings. Addressing these gaps with globally representative trials, standardized outcomes, and comprehensive implementation assessments will be critical to advancing diabetes prevention efforts and ensuring equitable access to effective interventions.

### Conclusions

This systematic review highlights the potential of both digital and in-person interventions for the prevention of T2DM, with digital interventions showing greater weight loss at 12 months but no significant differences at shorter or longer follow-up periods. However, the certainty of evidence was predominantly low to very low, and several prioritized key outcomes remain underexamined. The heterogeneity in intervention designs, lack of equivalence between modalities, and limited representation of diverse populations further underscore the need for caution when interpreting these findings. To strengthen the evidence base, future randomized controlled trials should prioritize equivalence in intervention intensity, longer follow-up durations, and the inclusion of key outcomes, such as cost-effectiveness, incidence of T2DM, and quality of life. These efforts are essential to guide the implementation of diabetes prevention programs and to ensure equitable access to effective interventions globally.

## Funding Statement

IRC is funded by Instituto de Salud Carlos III, grant number CP17/00017. RZC was funded by Instituto de Investigación Sanitaria de las Islas Baleares, grant number FOLIUM-2023 (founded by ITS2023/057). The rest of the authors are not funded by any grant or award to develop this work. The funders had no role in study design nor preparation of the protocol.

## Ethical approval

Not applicable (literature review)

## Data availability

The data that support the findings of this study are available from the corresponding author, IRC, upon reasonable request.

## Competing interests

The authors have no conflicts of interest to declare.

## Acknowledgments

We are grateful to the members of the international expert panel for the prioritization of the outcomes of this systematic review: Prof Miquel Benassar Veny (University of the Balearic Islands, Spain); Associate Professor Sonya Deschenes (University College Dublin, Ireland); Dr Jorge Caro (Instituto de Investigación Biomédica de Málaga, Spain), Dr Amy McInerney (University of Tübingen, Germany), Prof Pawel Lewek (Medical University of Lodz, Poland), Prof. Wayne Gao (Taipei Medical University, Taiwan), and Ana Sánchez (patient representative from the Spanish Federation of Diabetes).

## Online appendix 1. Search strategy

| N | Query | Results |
| --- | --- | --- |
| 1 | ((((((((exp Prevention/ or exp Primary Prevention/ or exp Risk Reduction Behavior/ or prevent*.mp. or prevention.mp. or "risk reduc*".mp. or "risk minimiz*".mp. or "risk control".mp. or prophylaxis.mp. or "health protect*".mp. or preventive.mp.) adj3 (strateg* or measur* or action* or intervent*).mp.) or prevent*.mp.) adj3 (strateg* or measur* or action* or intervent*).mp.) or risk.mp.) adj3 (reduc* or control or minimiz*).mp.) or early.mp.) adj3 intervent*.mp.) or disease.mp.) adj3 prevent*.mp. [mp=title, book title, abstract, original title, name of substance word, subject heading word, floating sub-heading word, keyword heading word, organism supplementary concept word, protocol supplementary concept word, rare disease supplementary concept word, unique identifier, synonyms, population supplementary concept word, anatomy supplementary concept word] | 219,383 |
| 2 | ((((((((((((((((exp Prediabetic State/ or exp Hyperglycemia/ or exp Glucose Intolerance/ or exp Insulin Resistance/ or exp Metabolic Syndrome/ or exp Prevention Programs/ or prediabet*.mp. or pre diabet*.mp. or "prediabetic state".mp. or "pre-diabetic state".mp. or "pre diabetes".mp. or "pre-diabetes".mp. or "impaired glucose tolerance".mp. or "glucose intolerance".mp. or hyperglycem*.mp. or "elevated blood glucose".mp. or dysglycem*.mp. or "impaired fasting glucose".mp. or "insulin resistance".mp. or "reduced insulin sensitivity".mp. or "high risk".mp. or risk.mp.) adj3 (diabet* or hyperglycem* or dysglycem* or "glucose intolerance").mp.) or "risk".mp.) adj3 diabet*.mp.) or prediabet*.mp.) adj3 (state or diagnos* or risk or factor*).mp.) or "glucose intolerance".mp.) adj3 (risk or factor* or diagnos*).mp.) or hyperglycem*.mp.) adj3 (risk or diagnos* or factor*).mp.) or "insulin resistance".mp.) adj3 (risk or diagnos* or factor*).mp.) or "metabolic syndrome".mp.) adj3 (risk or diagnos*).mp.) or "Diabetes Prevention Program".mp. or "DPP program".mp. or "Diabetes Prevention Study".mp. or "Diabetes Prevention Trial".mp. or "DPP intervention*".mp. or "DPP group*".mp. or "prevention program*".mp.) adj3 (diabetes or "risk reduc*" or "risk factor*").mp.) or "DPP".mp.) adj3 (intervention* or program* or trial* or group*).mp. [mp=title, book title, abstract, original title, name of substance word, subject heading word, floating sub-heading word, keyword heading word, organism supplementary concept word, protocol supplementary concept word, rare disease supplementary concept word, unique identifier, synonyms, population supplementary concept word, anatomy supplementary concept word] | 6,244 |
| 3 | *Diabetes mellitus/ or diabet*.ti,ab. or exp Diabetes Mellitus, Type 2/ or ((type 2 or type ii) adj2 diabet*).ti,ab. or ((non insulin* depend* or non insulin* depend* or non-insulin?depend* or non insulin?depend*) adj2 diabet*).ti,ab. or ((matur* onset or adult onset) adj2 diabet*).ti,ab. or (T2DM or T2D or TIIDM or TIID or NIDDM or MODY or MODM or AODM).ti,ab. or ((obes* or overweight) adj5 diabet*).ti,ab. | 586,736 |
| 4 | 1 and 3 | 10,374 |
| 5 | 2 or 4 | 15915 |
| 6 | Telemedicine/ or (mobile health or mhealth* or m-health or telehealth or tele-health or ehealth* or e-health* or (electronic adj2 health) or telemedicine).ti,ab. or (cell phones/ or (phone* or telephone* or smartphone* or smart-phone* or iphone* or i-phone*).ti,ab. or computers, handheld/ or (((computer* or device* or | 798,280 |
|  | technolog* or app*) adj4 (handheld* or hand-held* or palm* or mobile* or tablet* or pocket* or portable*)) or (PDA* or (Personal adj digital adj assistant*)))ti,ab.) or (text messaging/ or (((text* or short* or automat* or electron*) adj6 messag*) or sms).ti,ab. or Electronic mail/ or (email* or e-mail*).ab,ti. or ((internet* or www* or web* or computer* or net* or online* or on-line* or electronic* or automat*) adj5 (messag* or communicat*)))ti,ab.) or (digital* or online*).ti,ab. |  |
| 7 | trial*.ti,ab. or (randomized controlled trial or controlled clinical trial).pt. or ((cluster* or crossover* or quasiexperiment* or quasi-experiment* or control* or pragmatic) adj5 (trial* or study* or design*)))ti,ab. or (random* or (control* adj2 group*)))ti,ab. or RCT.ti,ab. | 3,234,548 |
| 8 | 5 and 6 and 7 | 510 |
| 9 | Meta-Analysis/ or Systematic Review/ or (review, systematic or systematic review or umbrella review).mp. or systematic review.ti. or narrative review.ti. | 480,908 |
| 10 | exp animals/ not humans.sh. | 5,290,921 |
| 11 | (rat or rats or rodent? or mice or mouse or cow or cows or cattle or calf or calves or ewe? or sheep or goat or ruminant? or chicken? or horse or horses or murine or bovine or ovine or animal?).ti. | 2,073,216 |
| 12 | 8 not (9 or 10 or 11) | 469 |

EMBASE (inception to 20 December 2024)
| No. | Query | Results |
| --- | --- | --- |
| #13 | #12 AND ('Article'/it OR 'Article in Press'/it) | 450 |
| #12 | #11 AND [embase]/lim NOT ([embase]/lim AND [medline]/lim) | 1,972 |
| #11 | #10 NOT (#6 OR #7) | 4,331 |
| #10 | #4 AND #5 AND #9 | 5,090 |
| #9 | #2 OR #8 | 757,389 |
| #8 | #1 AND #3 | 314,133 |
| #7 | 'animals'/exp NOT 'humans'/exp OR rat:ti OR rats:ti OR rodent*:ti OR mice:ti OR mouse:ti OR cow:ti OR cows:ti OR cattle:ti OR calf:ti OR calves:ti OR ewe*:ti OR sheep:ti OR goat:ti OR ruminant*:ti OR chicken*:ti OR horse:ti OR horses:ti OR murine:ti OR bovine:ti OR ovine:ti OR animal*:ti | 6,634,047 |
| #6 | 'meta-analysis'/exp OR 'systematic review'/exp OR 'review, systematic' OR 'systematic review' OR 'umbrella review' OR 'systematic review':ti OR 'narrative review':ti | 745,929 |
| #5 | trial*:ti,ab OR term:it OR (((cluster* OR crossover* OR quasiexperiment* OR 'quasi experiment*' OR control* OR pragmatic) NEAR/5 (trial* OR study* OR design*)))ti,ab) OR random*:ti,ab OR ((control* NEAR/2 group*)):ti,ab) OR rct:ti,ab | 4,304,118 |
| #4 | 'telemedicine'/exp OR 'mobile health':ti,ab OR mhealth*:ti,ab OR 'm health':ti,ab OR telehealth:ti,ab OR 'tele health':ti,ab OR ehealth*:ti,ab OR 'e health':ti,ab OR ((electronic NEAR/2 health):ti,ab) OR telemedicine:ti,ab OR 'cell phones'/exp OR phone*:ti,ab OR telephone*:ti,ab OR smartphone*:ti,ab OR 'smart phone':ti,ab OR iphone*:ti,ab OR 'i phone':ti,ab OR 'computers, handheld'/exp OR (((computer* OR device* OR technolog* OR app*) NEAR/4 (handheld* OR 'hand held*' OR palm* OR mobile* OR tablet* OR pocket* OR portable*)):ti,ab) OR pda*:ti,ab OR ((personal NEXT/1 digital NEXT/1 assistant*)):ti,ab) OR 'text messaging'/exp OR (((text* OR short* OR automat* OR electron*) NEAR/6 messag*)):ti,ab) OR sms:ti,ab OR | 1,131,206 |
|  | 'electronic mail'/exp OR email*:ti,ab OR 'e mail*:ti,ab OR (((internet* OR www* OR web* OR computer* OR net* OR online* OR 'on line*' OR electronic* OR automat*) NEAR/5 (messag* OR communicat*)):ti,ab) OR digital*:ti,ab OR online*:ti,ab |  |
| #3 | 'diabetes mellitus'/exp OR diabet*:ti OR 'diabetes mellitus, type 2'/exp OR (((('type 2' OR 'type ii') NEAR/2 diabet*):ti,ab) OR (((('non insulin* depend*' OR 'non-insulin\$depend*') NEAR/2 diabet*):ti,ab) OR (((('matur* onset' OR 'adult onset') NEAR/2 diabet*):ti,ab) OR t2dm:ti,ab OR t2d:ti,ab OR tiidm:ti,ab OR tiid:ti,ab OR niddm:ti,ab OR mody:ti,ab OR modm:ti,ab OR aodm:ti,ab OR (((obes* OR overweight) NEAR/5 diabet*):ti,ab) | 1,465,813 |
| #2 | 'prediabetic state'/exp OR 'hyperglycemia'/exp OR 'glucose intolerance'/exp OR 'insulin resistance'/exp OR 'metabolic syndrome'/exp OR 'prevention programs' OR prediabet* OR 'pre diabet*' OR 'prediabetic state' OR 'pre-diabetic state' OR 'pre diabetes' OR 'impaired glucose tolerance' OR 'glucose intolerance' OR hyperglycem* OR 'elevated blood glucose' OR dysglycem* OR 'impaired fasting glucose' OR 'insulin resistance' OR 'reduced insulin sensitivity' OR ('high risk' NEAR/3 (diabet* OR hyperglycem* OR dysglycem* OR 'glucose intolerance')) OR (risk NEAR/3 diabet*) OR (prediabet* NEAR/3 (state OR diagnos* OR risk OR factor*)) OR ('glucose intolerance' NEAR/3 (risk OR factor* OR diagnos*)) OR (hyperglycem* NEAR/3 (risk OR diagnos* OR factor*)) OR ('insulin resistance' NEAR/3 (risk OR diagnos* OR factor*)) OR ('metabolic syndrome' NEAR/3 (risk OR diagnos*)) OR 'diabetes prevention program' OR 'dpp program' OR 'diabetes prevention study' OR 'diabetes prevention trial' OR 'dpp intervention*' OR 'dpp group*' OR ('prevention program*' NEAR/3 (diabetes OR 'risk reduc*' OR 'risk factor*)) OR (dpp NEAR/3 (intervention* OR program* OR trial* OR group*)) | 527,937 |
| #1 | 'prevention'/exp OR 'primary prevention'/exp OR 'risk reduction behavior'/exp OR prevent* OR prevention OR 'risk reduc*' OR 'risk minimiz*' OR 'risk control' OR prophylaxis OR 'health protect*' OR (preventive NEAR/3 (strateg* OR measur* OR action* OR intervent*)) OR (prevent* NEAR/3 (strateg* OR measur* OR action* OR intervent*)) OR (risk NEAR/3 (reduc* OR control OR minimiz*)) OR (early NEAR/3 intervent*) OR (disease NEAR/3 prevent*) | 5,472,379 |

Cochrane Library (inception to 20 December 2024)
| ID | Search | Hits |
| --- | --- | --- |
| #1 | ((diabetes NEXT mellitus) OR diabet* OR (type NEXT 2 NEXT diabetes) OR ((type NEXT 2 OR type NEXT ii) NEAR/2 diabet*) OR ((non NEXT insulin* NEXT depend* OR non-insulin NEXT depend*) NEAR/2 diabet*) OR ((matur* NEXT onset OR adult NEXT onset) NEAR/2 diabet*) OR (T2DM OR T2D OR TIIDM OR TIID OR NIDDM OR MODY OR MODM OR AODM)) | 133,914 |
| #2 | ((prevent* NEAR/3 (strateg* OR measur* OR action* OR intervent*)) OR (prophylaxis NEAR/3 diabetes) OR ((disease NEAR/3 prevent*) AND (diabetes OR hyperglycem* OR glucose NEXT intolerance))) | 27,295 |
| #3 | #1 AND #2 | 4,844 |
| #4 | ((("prediabetic NEXT state") OR (hyperglycem* NEAR/3 diabetes) OR ("impaired NEXT glucose NEXT tolerance") OR ("insulin NEXT resistance") OR ("metabolic NEXT syndrome" AND diabetes) OR ("diabetes NEXT prevention NEXT program") OR (DPP NEAR/3 (intervention* OR program* OR group* OR trial*)) OR (DPP OR "diabetes NEXT prevention NEXT trial" OR "diabetes NEXT | 2,841 |
|  | prevention NEXT study")) |  |
| #5 | #3 OR #4 | 7,464 |
| #6 | (telemedicine OR (mobile NEXT health OR mhealth* OR m-health) OR (telehealth OR tele-health) OR (ehealth* OR e-health*) OR (mobile NEXT app* OR health NEXT app*) OR (app* NEAR/3 (health OR diabetes OR intervention*)) OR (smartphone NEXT app* OR phone NEXT app*) OR ((text NEXT messag* OR sms) OR ((short* OR automat*) NEAR/3 messag*)) OR (web-based NEXT intervention* OR internet NEXT intervention*) OR (online NEXT program* OR web NEXT platform*) OR ((digital NEXT tool* OR digital NEXT intervention*) NEAR/3 (diabetes OR health OR prevent*)) OR (wearable NEXT device* OR wearable*) OR ((electronic NEXT health) NEAR/3 (record* OR system*)) OR (remote NEXT monitoring OR virtual NEXT care OR remote NEXT consult*)) | 66,061 |
| #7 | #5 AND #6 | 1,131 |
|  | #7 AND TRIALS | 512 |

## Online appendix 2. List of outcomes prioritized by the international expert panel*

**Online appendix 2.** List of outcomes prioritized by the international expert panel\*
| **Outcome** | **Mean score** | **Prioritization** |
| --- | --- | --- |
| Change in BMI | 10 | Critical |
| Changes in HbA1c | 9 | Critical |
| Change in quality of life (e.g., EQ-5D-5L) | 9 | Critical |
| Change in waist circumference | 8 | Important |
| Change in physical activity level | 8 | Important |
| Change in body weight | 8 | Important |
| Change in diet | 8 | Important |
| Proportion of participants who develops T2DM based on WHO criteria | 7 | Important |
| Change in plasma glucose levels (2-hour oral glucose tolerance tests) | 7 | Important |
| Change in objectively measured sedentary behaviour: Inclinometer (eg, activPAL) or Accelerometer (eg, Actigraph, GENEActiv, Axivity) | 7 | Important |
| Change in self-reported smoking behaviour | 7 | Important |
| Cost-effectiveness (use of health-care services assessed by questionnaire and registers (associated costs calculated based on these data)) | 7 | Important |
| Change in resting blood pressure (resting systolic and diastolic blood pressure is measured by automatic sphygmomanometer in sitting position) | 6 | Not important |
| Change in objectively measured sleeping behaviour (e.g., Actigraph) | 6 | Not important |
| Change in self-reported alcohol consumption | 6 | Not important |
| Change in self-reported mental wellbeing | 6 | Not important |
| Work ability and associated costs (work ability is assessed by questionnaire; associated costs are calculated based on these data) | 6 | Not important |
| Change in plasma lipid concentration (plasma total, HDL and LDL cholesterol, and triglyceride concentrations are measured) | 5 | Not important |
| Change in cardiovascular disease risk (REGICOR) | 5 | Not important |
| Change in self-reported sedentary behaviour (change in sedentary behaviour is described as the total amount spent in sitting and lying position during waking hours, and is assessed by the question... | 5 | Not important |
| Change in insulin concentration (2-hour oral glucose tolerance tests) | 4 | Not important |
| Change in self-reported sleeping behaviour (according to sleeping behaviour questionnaire) | 4 | Not important |
| Change in self-reported eating behaviour | 4 | Not important |
| Change in inflammation markers (serum high-sensitivity C-reactive protein (hs-CRP) and interleukine-1Ra (IL-1Ra) are measured as biomarkers of inflammation.) | 3 | Not important |
| Change in liver function (Plasma aspartate aminotransferase (AST) and alanine aminotransferase (ALT) are measured as biomarkers of liver function.) | 3 | Not important |

BMI: Body Mass Index; EQ-5D-5L: EuroQol 5-Dimension 5-Level; HDL: High-Density Lipoprotein; HbA1c: Hemoglobin A1c; LDL: Low-Density Lipoprotein; T2DM: Type 2 Diabetes Mellitus; WHO: World Health Organization.
\*Outcomes with a mean score > 6 (in a scale 0-10) were prioritized for the systematic review

## Online appendix 3. List of articles excluded after full text screening with reasons

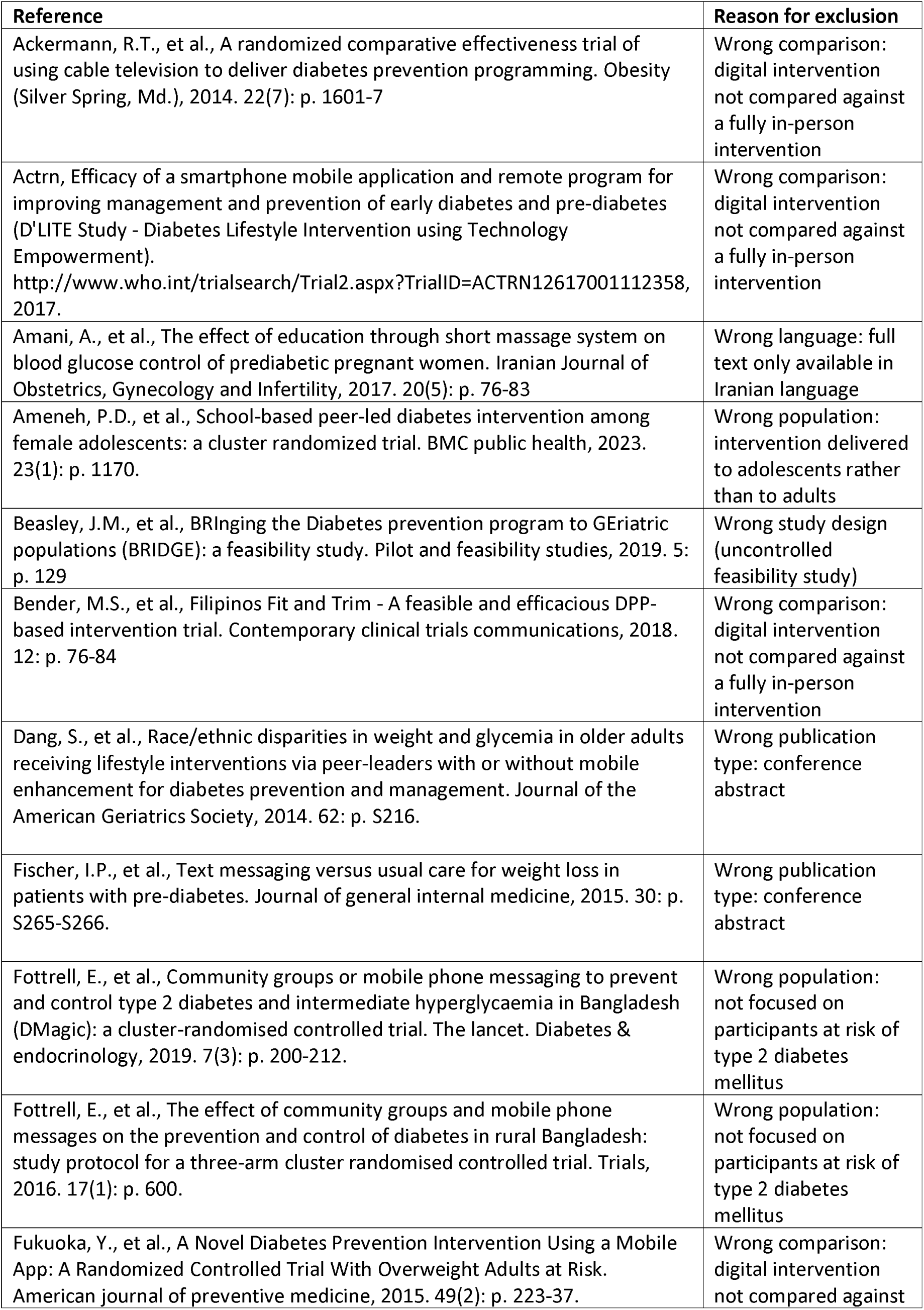
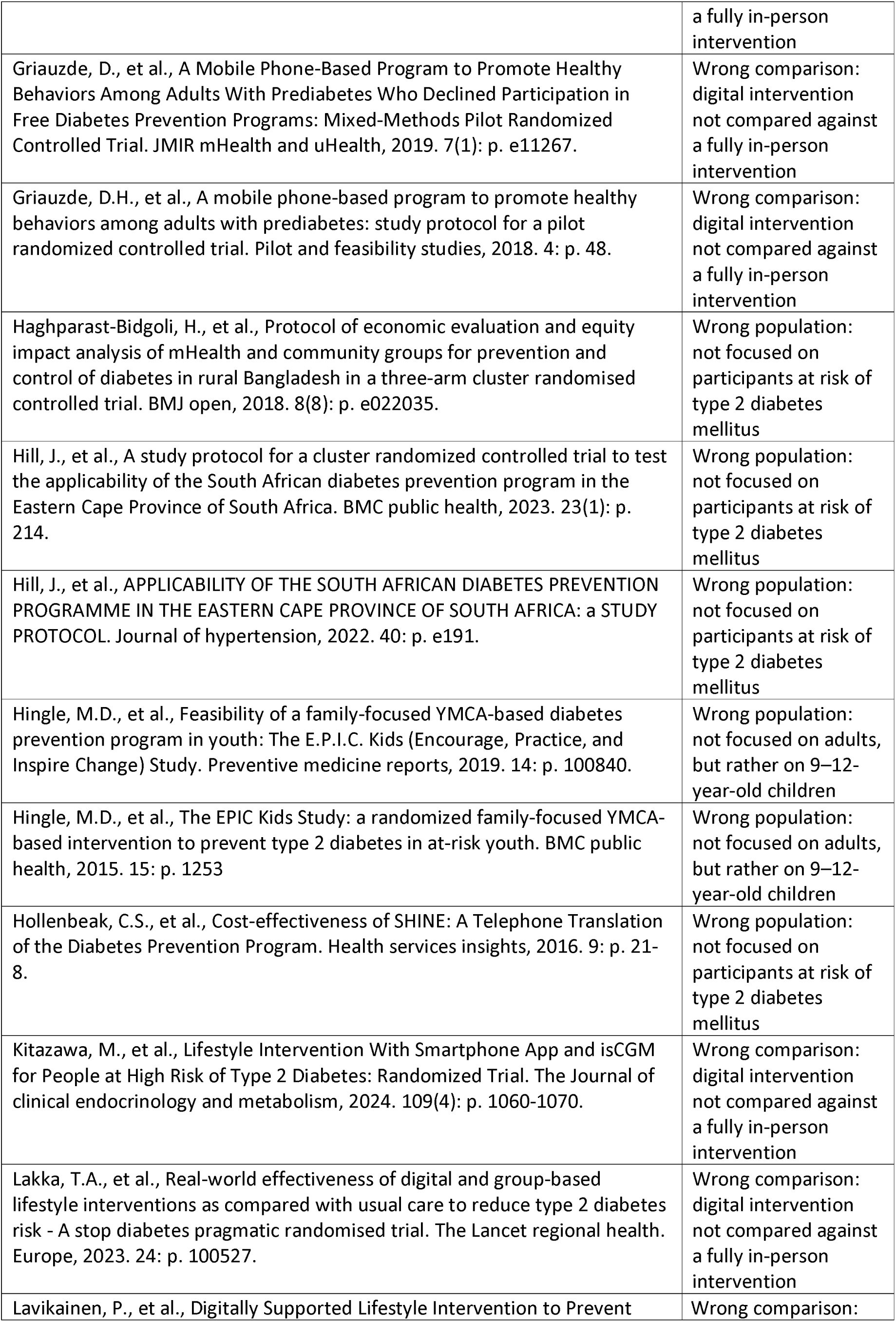
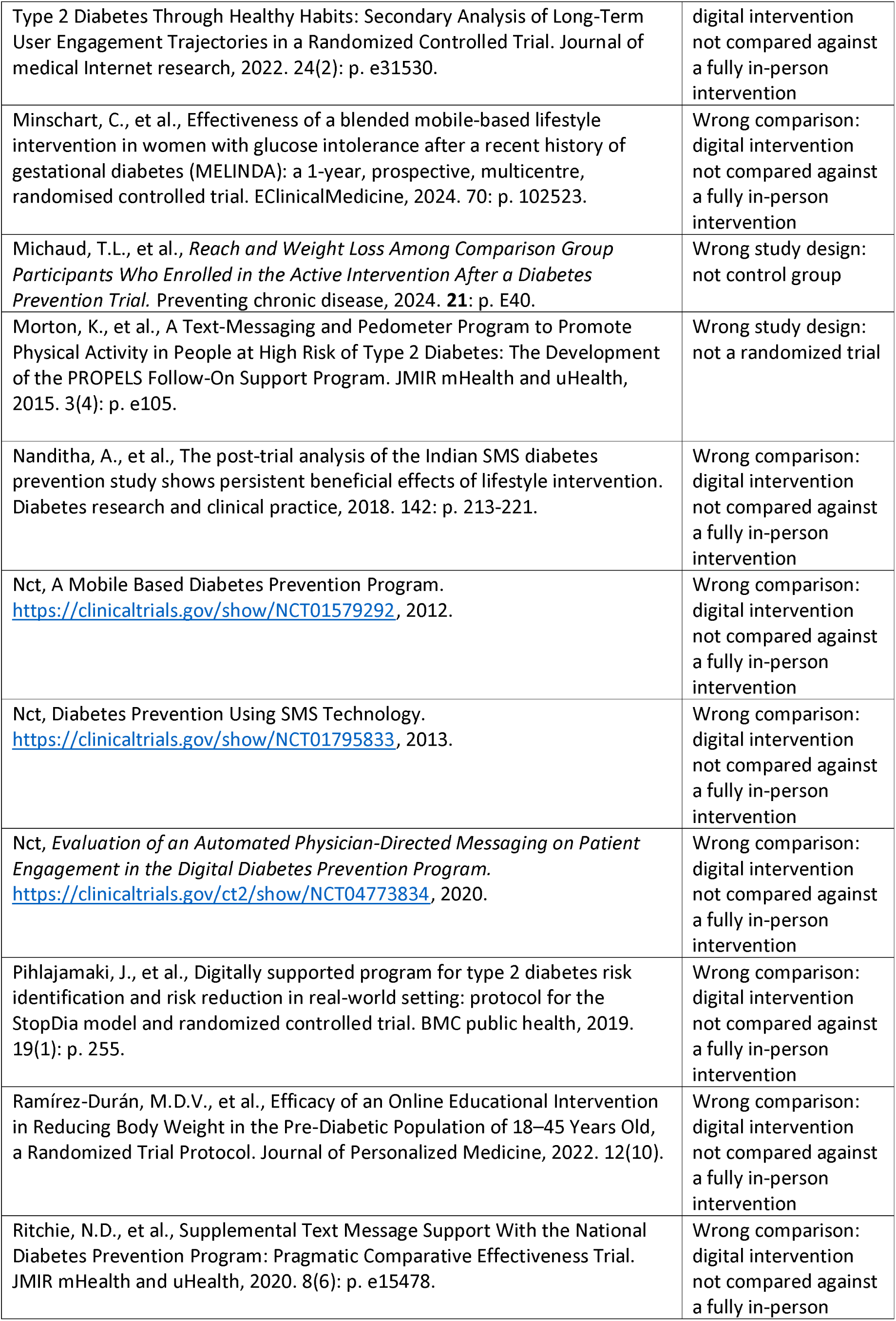
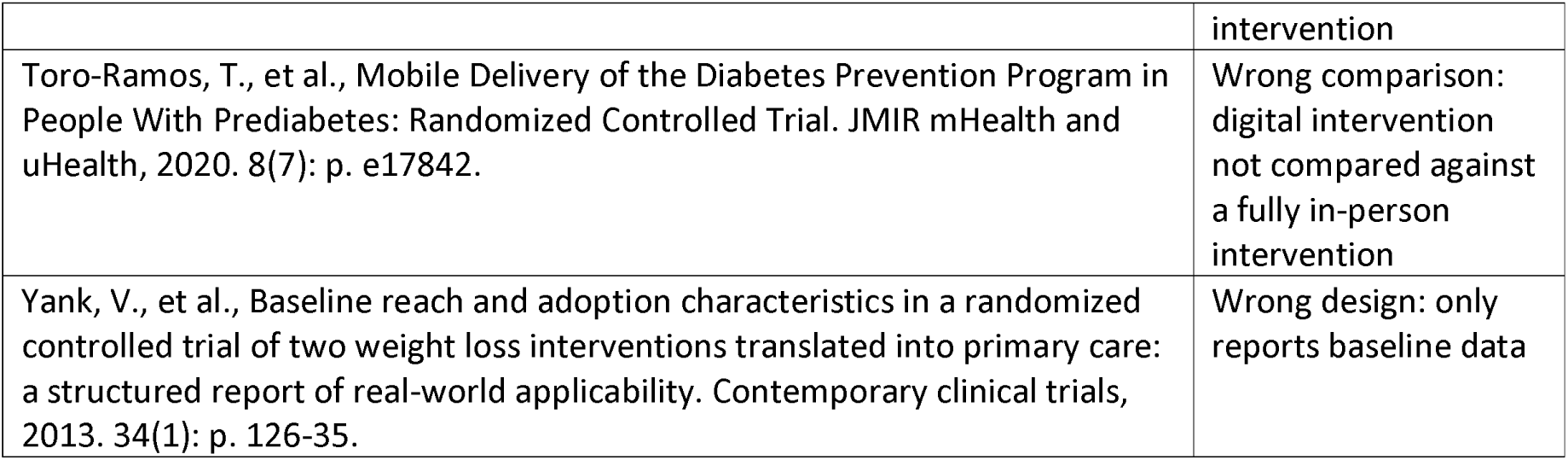

## Online Appendix 4. Characteristics of the ongoing trials identified

**Online Appendix 4.** Characteristics of the ongoing trials identified.
| **Trial name/**<br>**Trial code** | **Country** | **Trial design** | **Trial follow-up** | **Total number of participants** | **Age** | **Description of in-person intervention** | **N in-person intervention** | **Description of digital intervention** | **N digital intervention** |
| --- | --- | --- | --- | --- | --- | --- | --- | --- | --- |
| Abusamaan 2023[19]/<br>NCT05056376 | US | 12-month, parallel-group, non-inferiority, open-label, multicentre RCT | 6 and 12 months | 368 | 18–75 years | Human Coach-Based Diabetes Prevention Program: participants receive information about lifestyle change behaviors focusing on weight loss, physical activity, and nutrition from a trained lifestyle coach | 184 | Sweetch Digital Diabetes Prevention Program consists of a smartphone app and bluetooth-enabled digital body weight scale that syncs with the app. This app employs a sophisticated AI algorithm based on reinforcement learning | 184 |
| BRIDGE (Beasley 2023)[23]/<br>NCT05166785 | US | Type 1 hybrid, RCT | 6 and 12 months | 230 | ≥65 years | In-Person tailored for older adults DPP (DPP-TOAT): 22 intervention sessions over the course of a year by a certified DPP lifestyle coach | 115 | DPP tailored for older adults and delivered via Telehealth (V-DPP-TOAT): 22 intervention sessions over the course of a year by a certified DPP lifestyle coach | 115 |
| ai-DPP: Artificial Intelligence - powered Diabetes Prevention Program; NR: Not reported; RCT: Randomized Controlled Trial; US: United States |  |  |  |  |  |  |  |  |  |

## Online Appendix 5. Risk of bias of the included trials (detailed description)

|  | Randomization process |  | Deviation from intended interventions |  | Missing outcome data |  | Measurement of the outcome |  | Selection of the reported result |  | Overall risk of bias |  |
| --- | --- | --- | --- | --- | --- | --- | --- | --- | --- | --- | --- | --- |
| Trial name | Rating | Explanation | Rating | Explanation | Rating | Explanation | Rating | Explanation | Rating | Explanation | Rating | Explanation |
| PREDICTS | Low risk |  | Low risk |  | Low risk |  | Low risk |  | Low risk |  | Low risk |  |
| Moravcova 2024 | Some concerns | No specific information about the randomization process or allocation | Low risk | Although participants and researchers were aware of the intervention, no relevant deviations from protocol were reported, and ITT analysis conducted | Low risk | All documented reasons (Flow chart Fig 1) | Low risk | Assessment of the outcome "probably no" has been influenced by knowledge of intervention received | Low risk | Protocol available, 6-month follow up results reported (unique follow-up) | Some concerns | No data regarding randomization / allocation |
| Fuel Your Life | Some concerns | No allocation nor randomization methodology | Some concerns | No blinding information, and no trial reported and NI about deviations from the intended intervention | Some concerns | Maximum likelihood multiple imputation procedure was used (not appropriate sensibility analysis); no specific reasons reported of dropouts | Low risk | Knowledge about outcome assessors about the intervention arm of the participants could not (or difficultly) influence the outcome measurement | Some concerns | No protocol available | High risk | Overall, this trial lacks specific information regarding prior intentions, randomization, blinding, and the allocation process. Additionally, the high level of missing data was not adequately addressed through a robust sensitivity analysis. |
| E-LITE | Some concerns | No specific allocation method process reported | Low risk | Participants and researchers not blinded, but no deviations from protocol, and ITT analysis | Low risk | Sensitivity analyses were performed with participants with study-measured values | Low risk | Trained research assistants were unaware of participants, and outcome were correctly assessed | Low risk | Protocol available, all follow-up results provided | Some concerns | No specific allocation method process reported |
| DiaBEAT-it | Low risk | Prior to the baseline visit, the project manager created sealed opaque envelopes with group assignment information according to the blocked (groups of 4) randomization table stratified by gender created by the study statistician to blind research staff to intervention assignment. | Low risk | Participants blinded. No information about blinding on intervention researchers. No deviations from reported and ITT analysis conducted. | Some concerns | Important missingness (>20%). Missing data was replaced data with their baseline value following the "Baseline Carried Forward approach"; no information reported about specific missing reasons | Low risk | Outcome measurements are probably appropriate, and outcome assessors were blinded | Low risk | protocol available, and all results from all follow-ups were reported | Some concerns | No sensibility analysis nor correct approach to handle high missingness |
| CCSF Diabetes Prevention Trial (Ferrara 2020) | Some concerns | No information about allocation sequence | Some concerns | No blinding process reported | Some Concerns | Multiple imputation data sets analysis without sensibility analysis of the missing data | Low risk | Outcome measurements are probably appropriate, and outcome assessors were blinded | Some concerns | No protocol found | Some concerns | Overall, this trial lack of information on blinding, allocation sequence. There is also a lack of information on the existence of a prior protocol that would allow us to verify whether the study has suffered relevant deviations from the initial approach. |

## Online appendix 6. Forest plots of the random effects meta-analysis

### 6.1. Comparison of the effects at three months follow-up of digital vs in person interventions to prevent type 2 diabetes mellitus

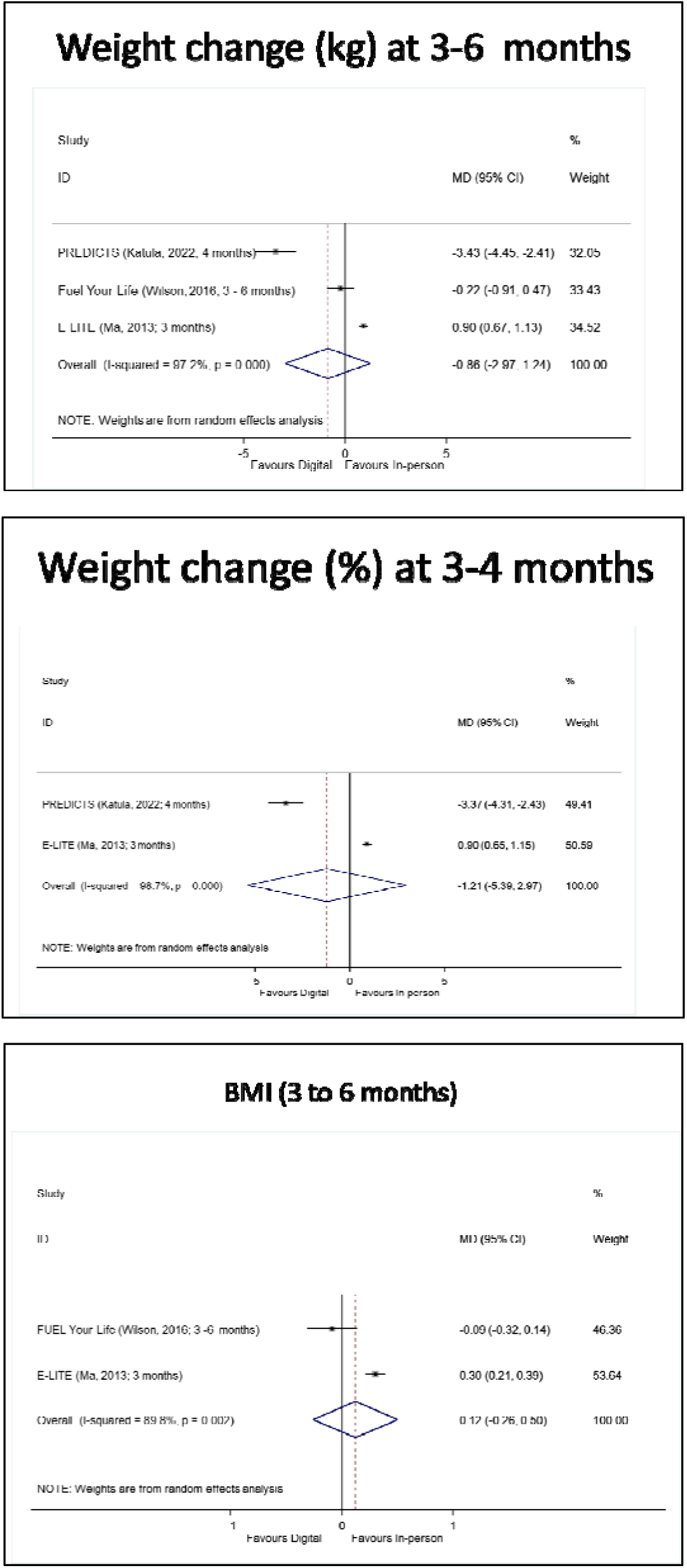

### 6.2. Comparison of the effects at six months follow-up of digital vs in person interventions to prevent type 2 diabetes mellitus

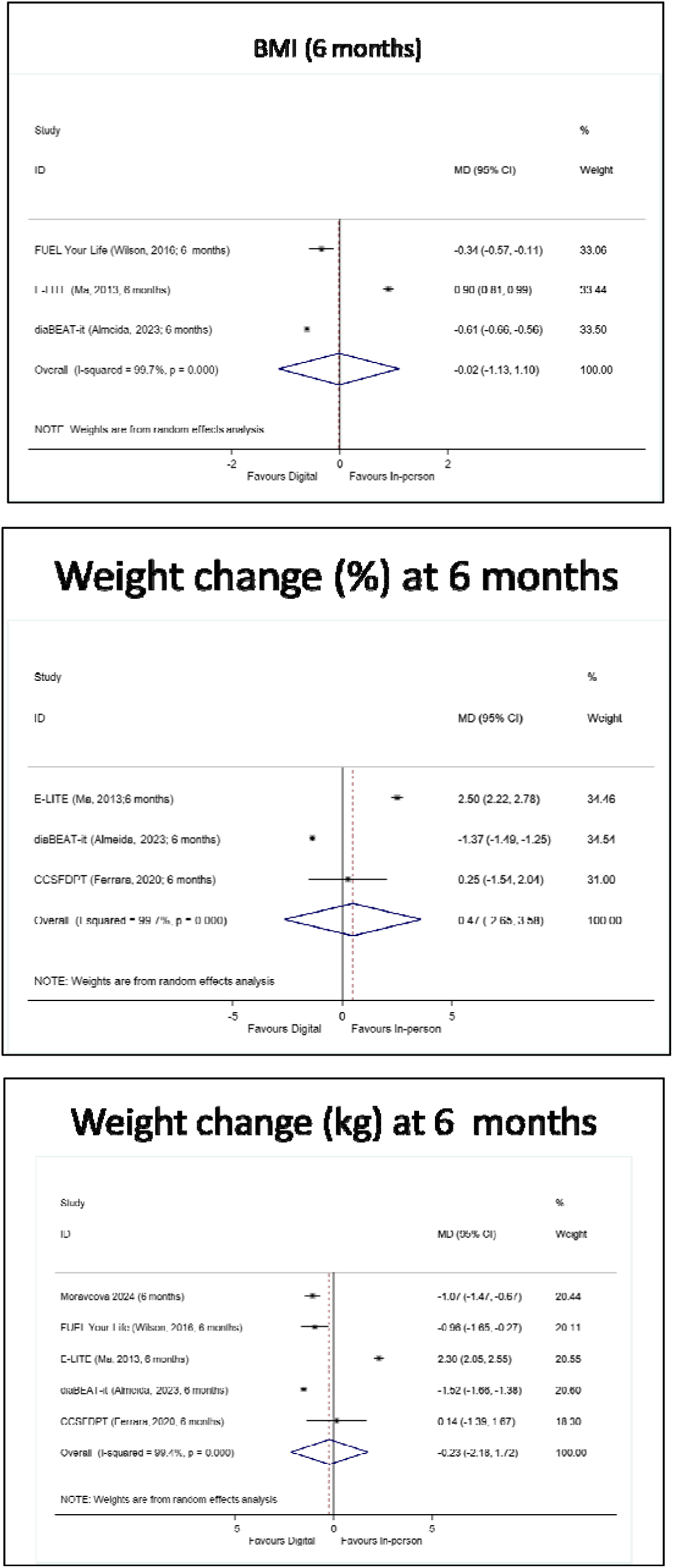

### 6.3 Comparison of the effects at 12 months follow-up of digital vs in person interventions to prevent type 2 diabetes mellitus

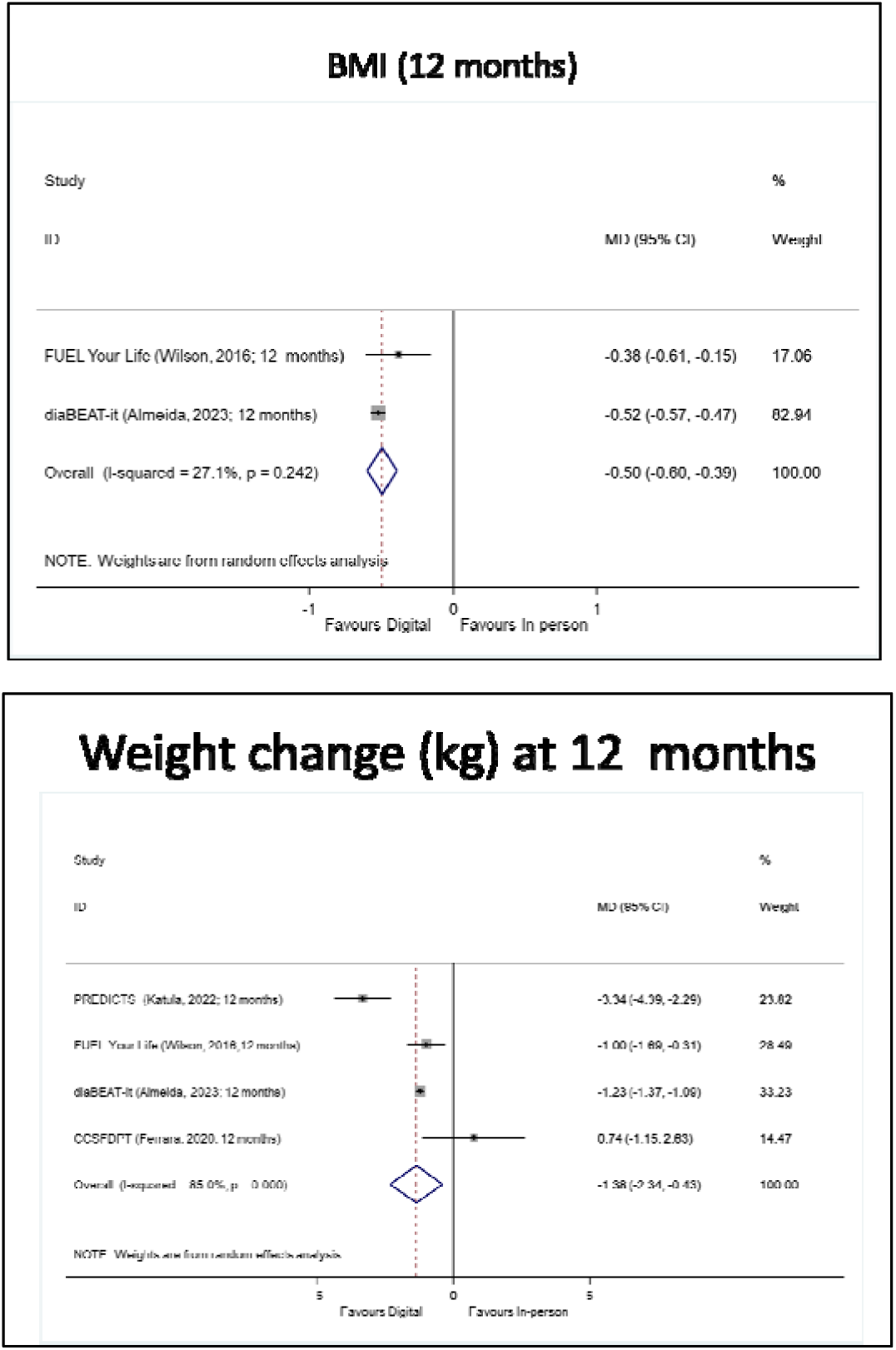

### 6.4. Comparison of the effects at >12 months follow-up of digital vs in person interventions to prevent type 2 diabetes mellitus

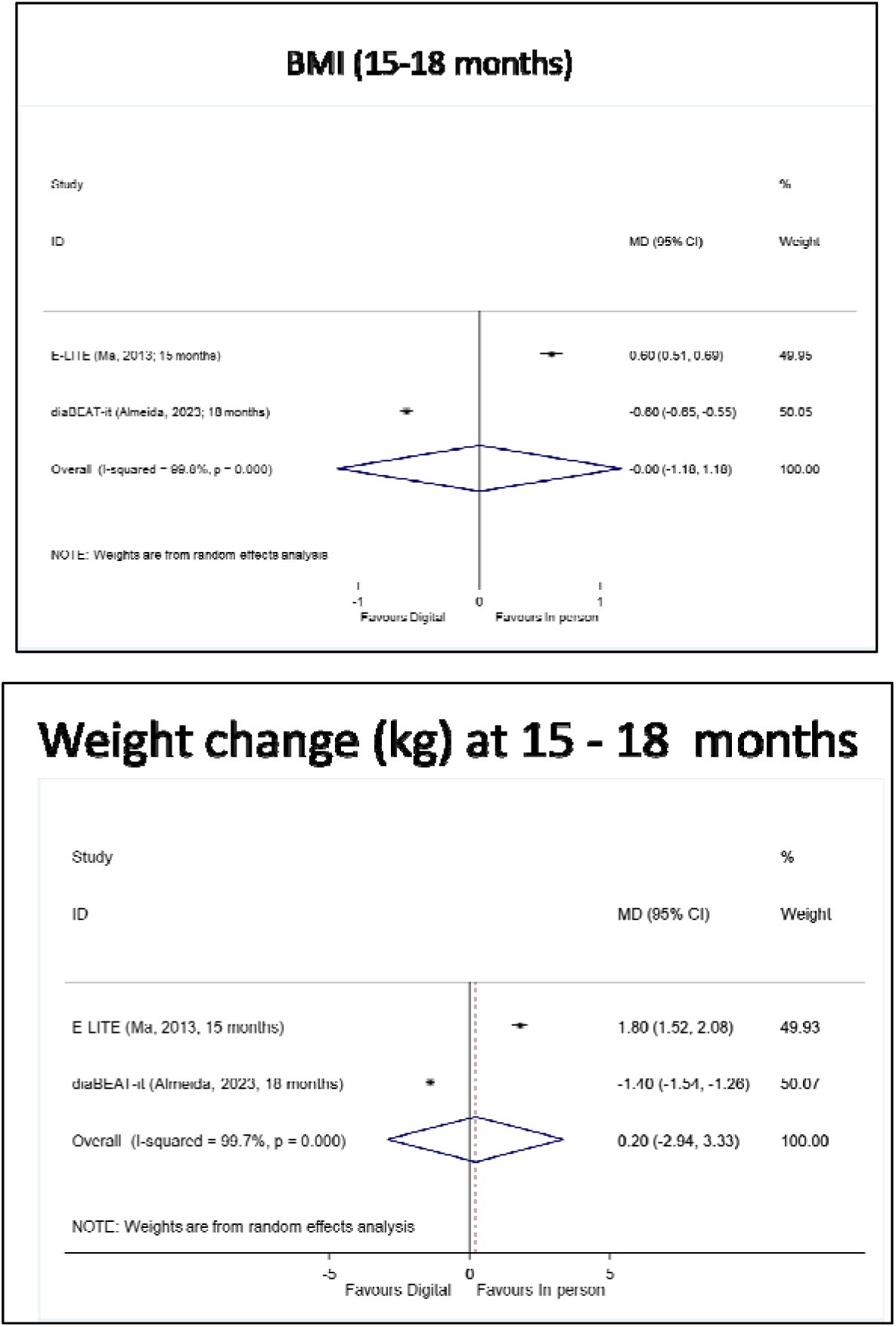

## Notes

### Competing Interest Statement

The authors have declared no competing interest.

### Funding Statement

IRC is funded by Instituto de Salud Carlos III, grant number CP17/00017. RZC was funded by Instituto de Investigacion Sanitaria de las Islas Baleares, grant number FOLIUM–2023 (founded by ITS2023/057). The rest of the authors are not funded by any grant or award to develop this work. The funders had no role in study design nor preparation of the protocol.

### Author Declarations

The study used openly available human data that were originally located at: 1.Abusamaan, M.S., et al., Effectiveness of artificial intelligence vs. human coaching in diabetes prevention: a study protocol for a randomized controlled trial. Trials, 2024. 25(1): p. 325. 2.Almeida, F.A., et al., Preventing diabetes with digital health and coaching for translation and scalability (PREDICTS): A type 1 hybrid effectiveness-implementation trial protocol. Contemporary clinical trials, 2020. 88: p. 105877. 3.Almeida, F.A., et al., Design and methods of "diaBEAT-it!": a hybrid preference/randomized control trial design using the RE-AIM framework. Contemporary clinical trials, 2014. 38(2): p. 383-96. 4.Almeida, F.A., et al., A randomized controlled trial to test the effectiveness of two technology-enhanced diabetes prevention programs in primary care: The DiaBEAT-it study. Frontiers in public health, 2023. 11: p. 1000162. 5.Beasley, J.M., et al., Study protocol: BRInging the Diabetes prevention program to GEriatric Populations. Frontiers in medicine, 2023. 10: p. 1144156. 6.Ferrara, A., et al., Comparative Effectiveness of 2 Diabetes Prevention Lifestyle Programs in the Workplace: The City and County of San Francisco Diabetes Prevention Trial. Preventing chronic disease, 2020. 17: p. E38. 7.Katula, J.A., et al., Effects of a Digital Diabetes Prevention Program: An RCT. American journal of preventive medicine, 2022. 62(4): p. 567-577. 8.Ma, J., et al., Translating the Diabetes Prevention Program lifestyle intervention for weight loss into primary care: a randomized trial. JAMA internal medicine, 2013. 173(2): p. 113-21. 9.Michaud, T.L., et al., Effects of a digital diabetes prevention program on cardiovascular risk among individuals with prediabetes. Primary care diabetes, 2023. 17(2): p. 148-154. 10.Michaud, T.L., et al., Cost and cost-effectiveness analysis of a digital diabetes prevention program: results from the PREDICTS trial. Translational behavioral medicine, 2023. 13(7): p. 501-510. 11.Moravcova, K., et al., Comparing the Efficacy of Digital and In-Person Weight Loss Interventions for Patients with Obesity and Glycemic Disorders: Evidence from a Randomized Non-Inferiority Trial. Nutrients, 2024. 16(10). 12.Nct, Preventing Diabetes With Digital Health and Coaching. https://clinicaltrials.gov/show/NCT03312764, 2017. 13.Nct, Effectiveness and Cost-Effectiveness of Fully-Automated Digital vs. Human Coach-Based Diabetes Prevention Programs. https://clinicaltrials.gov/ct2/show/NCT05056376, 2021. 14.Padilla, H.M., et al., Reach, Uptake, and Satisfaction of Three Delivery Modes of <ovid:i>FUEL Your Life</ovid:i>. Health promotion practice, 2021. 22(3): p. 415-422. 15.Park, S., et al., Cost-effectiveness analysis of a digital Diabetes Prevention Program (dDPP) in prediabetic patients. Journal of telemedicine and telecare, 2023: p. 1357633X231174262. 16.Wilson, M.G., et al., Effect of Intensity and Program Delivery on the Translation of Diabetes Prevention Program to Worksites: A Randomized Controlled Trial of Fuel Your Life. Journal of occupational and environmental medicine, 2016. 58(11): p. 1113-1120.

